# High variability in transmission of SARS-CoV-2 within households and implications for control

**DOI:** 10.1101/2021.01.29.20248797

**Authors:** Damon J.A. Toth, Alexander B. Beams, Lindsay T. Keegan, Yue Zhang, Tom Greene, Brian Orleans, Nathan Seegert, Adam Looney, Stephen C. Alder, Matthew H. Samore

## Abstract

**Background:** Severe acute respiratory syndrome coronavirus 2 (SARS-CoV-2) poses a high risk of transmission in close-contact indoor settings, which may include households. Prior studies have found a wide range of household secondary attack rates and may contain biases due to simplifying assumptions about transmission variability and test accuracy.

**Methods:** We compiled serological SARS-CoV-2 antibody test data and prior SARS-CoV-2 test reporting from members of 9,224 Utah households. We paired these data with a probabilistic model of household importation and transmission. We calculated a maximum likelihood estimate of the importation probability, mean and variability of household transmission probability, and sensitivity and specificity of test data. Given our household transmission estimates, we estimated the threshold of non-household transmission required for epidemic growth in the population.

**Results:** We estimated that individuals in our study households had a 0.41% (95% CI 0.32% – 0.51%) chance of acquiring SARS-CoV-2 infection outside their household. Our household secondary attack rate estimate was 36% (27% – 48%), substantially higher than the crude estimate of 16% unadjusted for imperfect serological test specificity and other factors. We found evidence for high variability in individual transmissibility, with higher probability of no transmissions or many transmissions compared to standard models. With household transmission at our estimates, the average number of non-household transmissions per case must be kept below 0.41 (0.33 – 0.52) to avoid continued growth of the pandemic in Utah.

**Conclusions:** Our findings suggest that crude estimates of household secondary attack rate based on serology data without accounting for false positive tests may underestimate the true average transmissibility, even when test specificity is high. Our finding of potential high variability (overdispersion) in transmissibility of infected individuals is consistent with characterizing SARS-CoV-2 transmission being largely driven by superspreading from a minority of infected individuals. Mitigation efforts targeting large households and other locations where many people congregate indoors might curb continued spread of the virus.

## 1 Introduction

Since its emergence in 2019, severe acute respiratory syndrome coronavirus 2 (SARS-CoV-2), the virus responsible for COVID-19, has spread rapidly, causing severe morbidity, mortality, and disruption to daily life. As public health officials continue grappling with reducing community spread, it is of increased importance to understand transmission risk in different locations where people mix. Transmission within households may be especially important, given the mounting evidence that indoor environments with close, sustained contact are especially high risk for SARS-CoV-2 transmission [1-3]. Furthermore, with substantial observed decreases in mobility during the pandemic [4], individuals likely are spending a greater proportion of time at home, thus increasing the importance of understanding within-household transmission. Likewise, isolation and quarantine measures recommended to help control COVID-19 frequently occur within homes, increasing risk to susceptible household members [5].

Data collected from members of households with at least one person infected with SARS-CoV-2 have revealed a wide range of within-household transmission estimates. One systematic review and meta-analysis [6] found 24 studies with household data conducted from January-March 2020, mostly in China, with secondary attack rate estimates ranging from 5% to 90% in the individual studies; pooling these data led to an average secondary attack rate estimate of 27% (95% CI: 21% – 32%). Another published review and meta-analysis of more recent data found 22 studies on the secondary attack rate in households, including estimates ranging from 4% to 32% [7]. Pooling these studies, the review found an average secondary attack rate of 17.1% (95% CI: 13.7% – 21.2%). Another review and meta-analysis found 40 household studies with individual study estimates ranging from 4% to 45% [8]. Their pooled analysis found that the household-based secondary attack rate for all household contacts was 19.0% (95% CI: 14.9 – 23.1%). Data from households in the U.S. [9-12] produced secondary attack rate estimates from 11% to 53%.

Most household studies generated data by first identifying index household cases via active or passive surveillance followed by monitoring and testing specimens from their household contacts using PCR or other methods that detect presence of the virus. These studies may exhibit bias if mild or asymptomatic cases were less likely to be identified as an index household case. By contrast, data for the presence of antibodies among household members provide information on the distribution of final sizes of household outbreaks no longer in progress and in which some or none of the cases were identified at the time. We are aware of only 3 studies that used serological antibody data to estimate household transmission, using data from Spain [13], Brazil [14], and Switzerland [15].

In addition to average transmission rates, heterogeneity and variability in SARS-CoV-2 transmission have also been quantified. The amount of individual-level variation in the number of secondary infections can affect final outbreak size [16]. Large variation (i.e., overdispersion) indicates the presence of superspreading by a minority of individuals who transmit to a disproportionately large number of others [17]. Better understanding of superspreading individuals and locations can greatly enhance efficient targeting of transmission control strategies [18]. Backward contact tracing can efficiently trace sources of acquisition to high-transmission individuals and circumstances when superspreading is present [19], and efforts that target similar circumstances for transmission prevention can have disproportionate benefits [20, 21].

Studies have quantified the variability in the number of SARS-CoV-2 transmissions from infected individuals using the dispersion parameter *k*, governing the variance of a negative binomially distributed offspring distribution [22-26]. Those studies estimated high overdispersion (low values of *k*) similar to what was observed during the first SARS-CoV outbreak in 2003 [17]. These estimates were derived from data on transmissions, including superspreading events, occurring in a variety of locations both inside and outside of households. Regarding household transmission specifically, Madewell et al. [8] showed preliminary evidence of overdispersion in household data, with more households than expected experiencing extremes of transmission (i.e., either no transmission or many transmissions) from an introduced case.

In this study, we combine SARS-CoV-2 data from serological antibody tests and self-reported prior tests to estimate within-household transmission of COVID-19 in Utah. Previously published secondary attack rate estimates are largely based on crude formulae which ignore the probabilities of multiple members of a household acquiring infection from the community, multiple generations of transmission within the household (i.e. secondary, tertiary, etc. transmissions), and imperfect test sensitivity and specificity. We addressed these limitations by extending previous models of final household outbreak size distributions [27] to develop a novel probabilistic model of household importation and household transmission combined with test sensitivity and specificity. Our model also quantifies variability in household transmission and the potential extent of overdispersion, to shed light on superspreading phenomena and the implications of household transmission for population-level controllability of COVID-19.

## 2 Methods

### 2.1 Data collection from Utah households

Details of our data collection process are described elsewhere [28]. Briefly, the Utah Health & Economic Recovery Outreach project involved selecting households in several counties in Utah by population sampling designed to form a set of households by which average community seroprevalence could be assessed. Any member of selected households could participate in a survey that included questions about prior SARS-CoV-2 test results (see Supplementary Methods for wording of relevant survey questions). Adult household members could fill out surveys on behalf of children of any age in the household. Survey participants age 12 or older could additionally opt to provide serological samples for COVID-19 antibody testing. Serum specimens were analyzed using the Abbott SARS-CoV-2 IgG assay performed on an Abbott Architect i2000 instrument (Abbott Laboratories), with methodology and criteria for a positive antibody result defined according to the manufacturer’s instructions. Data included in this analysis were collected between May 4 and August 15, 2020.

The University of Utah Institutional Review Board reviewed the surveillance project that produced the data analyzed in this manuscript and determined it as non-research public health surveillance, waived the requirement for documented consent, and determined that use of these data for analysis to understand the dynamics of SARS-CoV-2 transmission was exempt from further review (IRB_00132598). Individuals were informed of the project procedures and that participation was voluntary. Participants provided their agreement to participate and were given the chance to opt out of having their data used for future research. The data were analyzed anonymously for this manuscript.

The data are represented as follows. For each household in the dataset, we captured the following 7 values from the data:

- *n*: total number of people in household
- *a*: number who were antibody tested
- *s*: number who responded to the survey but were not antibody tested
- *a*_*PP*_: number who reported a prior positive test result and received a positive antibody test
- *a*_*PN*_: number who reported a prior positive test result and received a negative antibody test
- *a*_*NP*_: number who reported no prior positive test result and received a positive antibody test
- *s*_*P*_: number who were surveyed, reported a prior positive test result, and did not receive an antibody test

Those surveyed participants who reported no prior positive test result includes both those who had never been tested and those who had been tested but received no positive results. We did not have sufficient information to properly distinguish those two groups, nor to determine the circumstances of any prior negative tests that might affect the inferred probability of true prior infection.

Each of the *C* unique combinations of the above 7 values found at least once in the dataset was indexed as a vector **y**_*i*_:

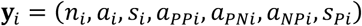

We tallied the number of households for which each **y**_*i*_ occurred in the frequency elements *f*_*i*_, and represented the entire dataset by the vector **y** = (**y**_1_, …, **y**_*C*_, *f*_1_, … *f*_*C*_).

The dataset **y** and all codes, written in R version 4.0.3, used for analyses described in the following sections are posted and publicly available at https://github.com/damontoth/householdTransmission.

### 2.2 Total household infection size model

Here we derive the probabilities *M*_*kn*_ for the probability that *k* out of *n* total household members ended up infected. If *k* members of a size-*n* household were infected, that means that *n* − *k* members escaped being infected by a non-household member (called “community” acquisitions) and escaped being infected by any of the *n* infected within the household. Thus, our model for *M*_*kn*_ combines both probabilities and does not depend on the order of occurrence of household transmissions and subsequent community acquisitions after the initial one, as in similar prior formulations [27]. Also following prior formulations, we assume that active infections were not present in the households at the time of antibody data collection (i.e., that household outbreaks had reached final size). Accounting for the timing of recent household importations, transmissions, and development of detectable antibodies during an ongoing household outbreak would significantly complicate the model equations and would likely have little effect on our overall results, given that the prevalence of active infections at the time of data collection was very low [28].

The *M*_*kn*_ values depend on 3 parameters. The parameter *p*_*c*_ is the average per-capita probability of community acquisition, *p*_*h*_ is the mean transmission probability from an infected person to a fellow household member, and *d*_*h*_ is the dispersion parameter characterizing variability in transmissibility across infected individuals, with no assumed correlation among members of the same household.

For a given household size *n* ≥ 2, the formula for *M*_*kn*_ is:

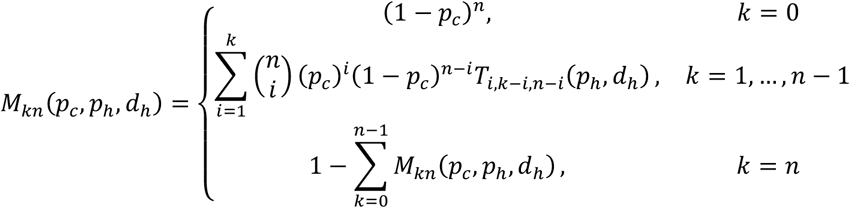

For households of size *n* = 1, note that the expression involving the household transmission parameters does not apply and we have *M*_01_ = 1 − *p*_*c*_ and *M*_11_ = *p*_*c*_.

The probability that a household of size *n* had 0 infections: *M*_0*n*_ = (1 − *p*_*c*_)^*n*^, is the probability that none of the household members acquired infection from the community and does not depend on the household transmission variables because no household transmissions were possible without a community introduction. For the final number of household infections to be nonzero, there must be at least one community acquisition, which may be followed by within-household transmissions. The 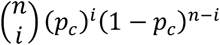 expression is the binomial probability that *i* out of the *n* household members had a community acquisition, and the function *T*_*xyz*_ is the probability that *x* already infected household members lead to a total of *y* transmissions to *z* susceptible household members. In other words, *T*_*xyz*_ is the probability that the final outbreak size is *x* + *y*, given that *x* household members are already infected in a house with *z* susceptible members. For efficiency of computation, the *T*_*xyz*_ values are calculated in order of increasing values of *y*, i.e. *T*_*x*0*z*_ for each relevant *x* and *z* value are calculated first, then the *T*_*x*1*z*_ values, then *T*_*x*2*z*_. This allows the use of *T*_*xyz*_ values for lower values of *y* to be used in the formula (see Supplementary Material for details):

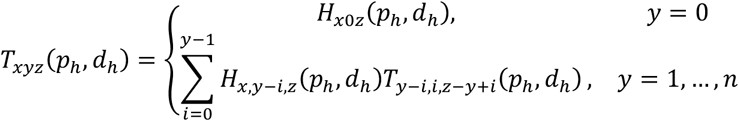

Within the *T*_*xyz*_ formula, the function *H*_*xyz*_ is the probability that *x* infected household members transmit infection *directly* to *y* out of *z* fellow household members who are susceptible. The *H*_*xyz*_ values are calculated in order of increasing values of *x* for efficient computation (see Supplementary Material):

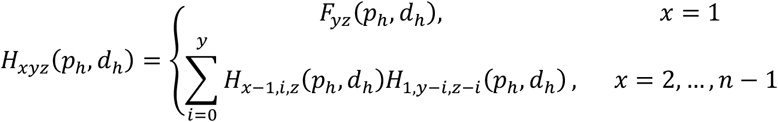

Finally, the function *F*_*yz*_(*p, d*) is the probability mass function of the beta-binomial distribution for *y* successes out of *z* trials, parameterized by a mean success probability *p* and a dispersion parameter *d*. When *d* is finite and nonzero, *F*_*yz*_ is derived from the binomial distribution with success probability that is a beta-distributed random variable with parameters *α* = *dp, β* = *d*(1 − *p*), with decreasing variance as *d* increases. We also make use of the boundary cases *d* = 0 and *d* → ∞. In the limit *d* → ∞, holding *p* constant, *F*_*yz*_ becomes the binomial distribution with constant success probability *p* (Supplementary Material). In the maximal variance limit, *d* → 0, with *p* held constant, *F*_*yz*_ becomes an “all-or-nothing” distribution where *y* = *z* successes occur with probability *p* and to *y* = 0 successes occur with probability 1 − *p* (Supplementary Material):

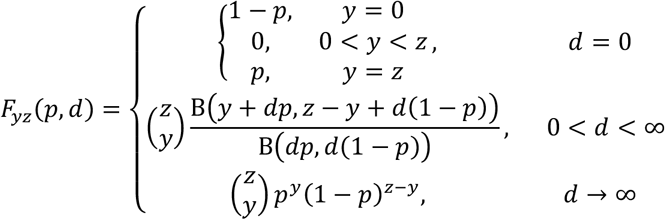

The function B is the beta function. We use *F*_*yz*_ within the formula for *H*_1*yz*_ to quantify the distribution of household transmissions directly from a single infected household member, where *y* is the number of transmissions, *z* is the number of susceptible household members, *p* = *p*_*h*_, and *d* = *d*_*h*_.

The above formulas are derived in the Supplementary Material. Elements of this model appear in other publications. Longini and Koopman [27] derived a formula for *M*_*kn*_ for the model with no variability among households or individuals, equivalent to our model with *d*_*h*_ → ∞. While they provided a more efficient formula that takes advantage of the properties of that special case, we confirmed that our calculation scheme above reproduces the results of their formula. Becker [29] published explicit formulas for the final size of household outbreaks after a single introduction to households up to size 5 using the beta-binomial chain model, equivalent to our *T*_*xyz*_ for *x* = 1 and *z* up to 4. We confirmed that our scheme for calculating *T*_*xyz*_ produces the same results as their example formulas for arbitrary values of *p*_*h*_ and *d*_*h*_.

### 2.3 Likelihood model

We sought to use our data to simultaneously estimate the 3 parameters (*p*_*c*_, *p*_*h*_, *d*_*h*_) using maximum likelihood estimation (MLE). However, applying the *M*_*kn*_ formula directly to our data would be problematic because the true number of infections *k* in each household are not known with certainty. The data include two sources of COVID-19 test information by which prior infection status of a portion of individual household members can be probabilistically inferred: antibody test results and surveys in which participants could report results of a priortest.

Antibody test results are subject to imperfect sensitivity and specificity due to false negative tests and false positive tests, respectively. To account for these, we added two additional parameters to be estimated by the MLE: *ϕ*_*A*_, the probability that an antibody-tested person with a prior infection tested positive for antibodies, and *π*_*A*_, the probability that an antibody-tested person with no prior infection tested negative for antibodies.

Prior test results for SARS-CoV-2 reported on the survey also do not perfectly identify those with prior infections. To quantify this imperfection, we introduced two more parameters to be estimated by the MLE: *ϕ*_*V*_, the probability that a surveyed person with a prior infection reported receiving a positive test for the virus, and *π*_*V*_, the probability that a surveyed person with no prior infection did not report receiving a positive test.

Some household members received a survey but no antibody test and other members received neither. The *M*_*kn*_ formula depends on the total household size *n*, which for many households includes individuals with missing data. For households with at least one but not all members infected (1 ≤ *k* ≤ *n* − 1) and in which less than *n* member were full participants, the likelihood formula required the probability that different portions of the *k* infected members were among those who were antibody tested or surveyed only. To arrive at our formula, we assumed that the antibody-tested and surveyed-only portion of a household were a random sample of household members with respect to their prior infection status. I.e., we assumed that those individuals in a participating household with and without prior infections were equally likely to participate in the study and equally likely to agree to antibody testing.

In all we have 7 variables to be estimated by MLE, encapsulated in the following vector **θ**:

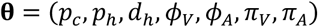

The log likelihood of the dataset **y** described in Section 2.1 with variable set **θ** is then

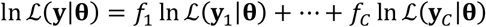

To present the formula for ℒ(**y**_*i*_|**θ**), the likelihood of a particular **y**_*i*_, we first define the following quantities calculated from the core elements of **y**_*i*_ listed in Section 2.1:

- *a*_*NNi*_ = *a*_*i*_ − *a*_*PPi*_ − *a*_*PNi*_ − *a*_*NPi*_: number who reported no prior positive test result and received a negative antibody test
- *s*_*Ni*_ = *s*_*i*_ − *s*_*Pi*_: number who were surveyed, reported no prior positive test result, and did not receive an antibody test
- *q*_*i*_ = *n*_*i*_ − *a*_*i*_ − *s*_*i*_: number untested for antibodies and not surveyed

Then we have:

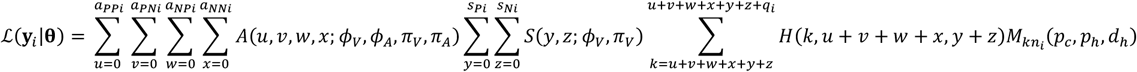

In the formula, the function *A* quantifies the probability of observing the given set of test result combinations among antibody-tested people (*a*_*PPi*_, *a*_*PNi*_, *a*_*NPi*_, *a*_*NNi*_), given that (*u, ν, w, x*) of them had a prior infection, respectively. E.g., *u* is the number of the *a*_*PPi*_ household member who had an infection (true positives), *ν* is the number of the *a*_*PNi*_ household members who had an infection (true positive by prior test and false negative by antibody test), *w* is the number of the *a*_*NPi*_ household members who had an infection (true positive by antibody test and did not report a prior positive test), and *x* is the number of the *a*_*NNi*_ household members who had an infection (false negative by antibody test and did not report a prior positive test). The formula for *A* is

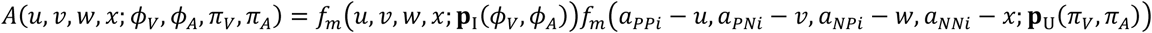

The function *f*_*m*_(**r**; **p**) is the probability mass function for the multinomial distribution, where the number of trials is the sum of the elements of **r**, which are the number of infected or uninfected antibody-tested people who received each of the four possible test result combinations. The vector **p** contains the probability of each of the four test result combinations given that the person was infected (for **p** = **p**_I_) or uninfected (for **p** = **p**_U_):

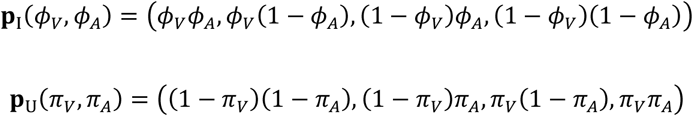

The first element of **p**_I_, *ϕ*_*V*_*ϕ*_*A*_, is the probability that an antibody-tested person with a prior infection reported a prior positive test (with probability *ϕ*_*V*_) and also had a positive antibody test result (with probability *ϕ*_*A*_). Note that *ϕ*_*A*_ represents the sensitivity of the antibody test, but *ϕ*_*V*_ includes both the sensitivity of the prior test and the probability that an infected person actually sought and received a SARS-CoV-2 test during the period of infection in which detectable virus was present and reported that positive test on our survey. Elements 2–4 of **p**_I_ are the probabilities that an antibody-tested, prior infected person reported a prior positive test but tested negative for antibodies, did not report a prior positive test and tested positive for antibodies, and did not report a prior positive test and tested negative for antibodies, respectively. The elements of **p**_U_ are the corresponding probabilities for individuals with no prior infection.

The function *S* quantifies the probability of the survey-only data (*s*_*Pi*_, *s*_*Ni*_) given that *y* of the *s*_*Pi*_ individuals had a prior infection and *z* of the *s*_*Ni*_ individuals had a prior infection:

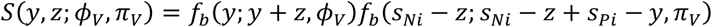

The function *f*_*b*_(*q*; *r, p*) is the probability mass function for the binomial distribution, for *q* successes given that there were *r* independent trials with probability *p* for success of each trial.

The function *H*(*k, k*_*a*_, *k*_*s*_) in the likelihood equation is the probability that, when *k* of *n*_*i*_ individuals in the household were infected, *k*_*a*_ infected individuals were among the *a*_*i*_ individuals antibody tested and *k*_*s*_ infected individuals were among the *s*_*i*_ individuals surveyed but not antibody tested:

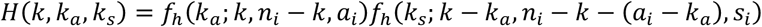

The function *f*_*h*_(*b*; *c, d, e*) is the probability mass function of the hypergeometric distribution for the number *b* of infected people selecting to be antibody-tested or surveyed-only, given that there were *c* infected people and *d* uninfected people available for selection in the household, and *e* people were tested or surveyed-only. These terms account for individuals in households who received neither an antibody test nor a survey, who may have included infected individuals. Our use of the hypergeometric distribution led from our assumption that, if some members of the household had a prior infection and others didn’t, the antibody-tested / surveyed individuals were a random sample from the household with respect to their prior infection status.

### 2.4 Likelihood optimization and uncertainty

We maximized the log likelihood over the 7 unknown parameters (*p*_*c*_, *p*_*h*_, *d*_*h*_, *ϕ*_*V*_, *ϕ*_*A*_, *π*_*V*_, *π*_*A*_) using the observations (*n, a, s, a*_*PP*_, *a*_*PN*_, *a*_*NP*_, *s*_*P*_) for each household, to produce the MLE: 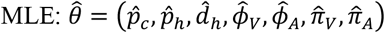. The log likelihood maximization was performed using the “optim” function in R. We derived approximate confidence interval boundaries for an individual parameter *θ*_*i*_ using the likelihood ratio test, using the statistic 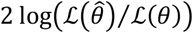, where *θ* consists of *θ*_*i*_ freely varying and the other 6 elements of *θ* held at their optimal value. We defined a 95% confidence interval boundary where *θ*_*i*_ produces a value for this statistic equal to the 95^th^ percentile of the chi-squared distribution with 1 degree of freedom. We also plotted 2-dimensional confidence region boundaries for each of the 21 possible (*θ*_*i*_, *θ*_*j*_) parameter pairs by allowing each pair to vary freely together while holding the other 5 at their optimal values. We calculated the boundary in the (*θ*_*i*_, *θ*_*j*_) parameter plane where the likelihood ratio statistic equals the 95^th^ percentile of the chi-squared distribution with 2 degrees of freedom. To calculate P-values at which certain fixed parameter values could be rejected in favor of the MLE, we used the chi-squared distribution with degrees of freedom equal to the number of fixed parameters.

Additionally, we developed a simulation model to produce synthetic data sets on which to test our likelihood model. We ran the simulation for the same number of households with the same sizes and participation rates for survey and antibody testing as in the actual data (fixed values of *n, a*, and *s* for each household). We randomized importations to households and simulated transmissions using the MLE values of the three epidemiological parameters *p*_*c*_, *p*_*h*_, and *d*_*h*_, randomized survey and antibody test results using the MLE sensitivity and specificity values, and maximized the likelihood against the simulated data. We repeated this process for 500 simulated data sets and recorded the median estimated value of each variable, for comparison against the MLE value that generated the data. We also used the 500 sets of simulation-based estimates as a parametric bootstrap to generate 95% confidence estimates for each variable, for comparison against the intervals generated from the likelihood ratio test.

Finally, we tested an alternate model that allows the community acquisition probability to vary by household, such that some households may have a higher per-capita acquisition rate than others applied to each household member. To quantify this probability in the alternate model, we employed the beta-binomial distribution for the number of community acquisitions in a household of a given size (see Supplementary Methods).

### 2.5 Household transmission variability

We quantified the implications of our household transmission variability estimates by calculating the probability of transmission extremes, compared to those produced by the classic binomial transmission model (*d*_*h*_ = ∞). Specifically, we calculated the probability that an initially infected individual transmits to no one or everyone in households of sizes from 2 to 10. For households of size *n*, the probability of no transmissions from the index infection is *F*_0,*n*−1_(*p*_*h*_, *d*_*h*_) and the probability the index person transmits directly to the entire household is *F*_*n*−1,*n*−1_(*p*_*h*_, *d*_*h*_). We used our overall MLE values for 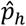 and 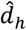 to calculate these values for each *n*, with confidence intervals using our parametric bootstrap results. For comparison to the binomial model we applied *d*_*h*_ = ∞, paired with the alternate MLE of *p*_*h*_ under that constraint.

We also calculated an example of a dynamic transmission model that produces a distribution of household transmission probabilities close to that produced by our MLE beta distribution, using the method of moments. Specifically, if an infected person’s duration of infectiousness is assumed to be fixed and transmissibility to a housemate is modeled as a gamma distribution with shape parameter *k*, then we solve for the value *k* that produces the same mean and variance for the transmission probability as that of the beta distribution with mean *p*_*h*_ and dispersion *d*_*h*_ (Supplementary Methods). We solved for *k* using our 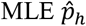 and 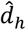 values, and we derived a confidence interval for *k* using the pairs of (*p*_*h*_, *d*_*h*_) estimates from our parametric bootstrap analysis.

### 2.6 Within-household reproduction number

We calculated the within-household reproduction number *R*_*h*_, defined as the expected number of household transmissions directly from a community acquirer with all fellow household members susceptible:

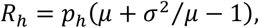

where *μ* and *σ*^2^ are the mean and variance of the household size distribution, and *p*_*h*_ is the secondary attack rate as determined by our MLE. This equation for *R*_*h*_ is derived as in Ball et al. [30] and detailed in the Supplementary Material.

Additionally, we derived an alternate household reproduction number 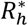 defined as the expected total number of transmissions in the household of an infected person who acquired infection in the community and has no initially non-susceptible housemates. This differs from *R*_*h*_ in that it counts all potential downstream transmissions in the household stemming from the index community acquirer. The formula for 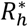, derived in the Supplementary Material, is

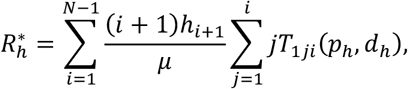

where *h*_*i*_ is the fraction of all households that are size *i*.

To investigate the implications of household transmission for population-wide transmission control, we use a threshold condition delineating subcritical and supercritical transmission in the population. Supercritical transmission occurs when 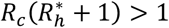, where *R*_*c*_ is the average number of community (non-household) transmissions from an infected person. We derive this formula in the Supplementary Material, following Ball et al. [30]. We estimated *R*_*h*_, 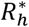, and the threshold value for *R*_*c*_ by applying our MLE estimates of *p*_*h*_ and *d*_*h*_ to the above formulas and their confidence intervals by applying the (*p*_*h*_, *d*_*h*_) pairs from each parametric bootstrap estimate.

## 3. Results

### 3.1 Data summary

We compiled data from 9,383 households (Figure 1). Of these, we retained 9,224 (98.3%) for use in the MLE. The 159 excluded households were removed because the household size was unknown (51) or the reported household size was less than the number of people tested or surveyed in the house (108). In the 9,224 retained households, there were 28,321 (3.07 per household) reported household members, 13,998 (1.52 per household) people who were both surveyed and antibody tested, and another 5,249 (0.57 per household) who were surveyed but not antibody tested. The households in the data were located in 7 of the 29 counties in Utah; the 22 excluded counties account for <14% of Utah’s total population Supplementary Table S1.

**Figure 1.**
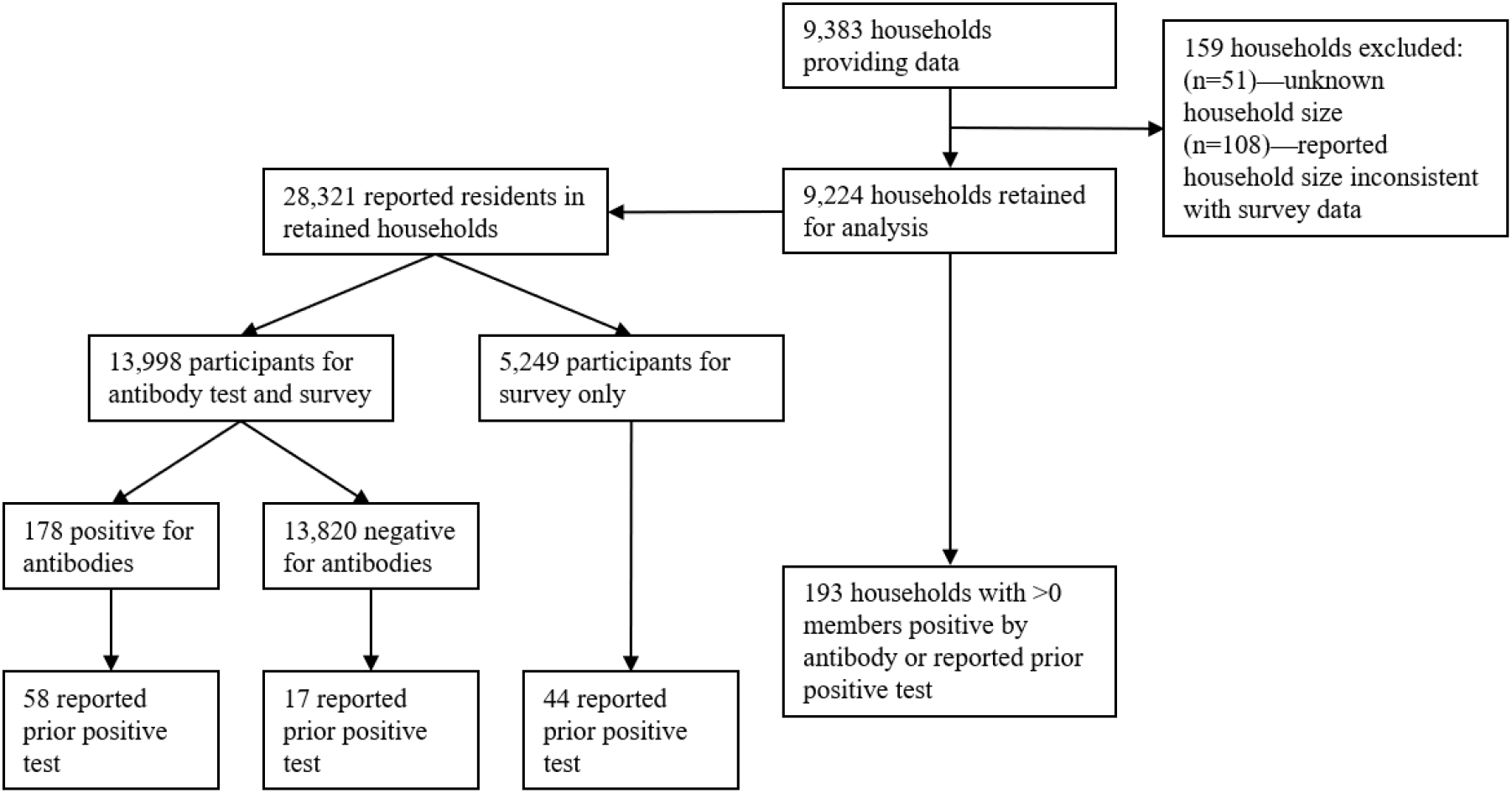
Data summary flowchart. Flow diagram for data from participating households and household members

Of the 13,998 antibody tests in the retained households, 178 (1.27%) were positive. Of those 178 people with a positive antibody test, 58 (32.6%) reported receiving a prior positive test. Of the 19,247 people who were antibody tested or surveyed only, 119 (0.62%) reported receiving a prior positive test. This broke down to 0.53% (75 / 13,998) for those who were antibody tested and 0.84% (44 / 5,249) for those who were surveyed but not antibody tested. The rate of testing positive for antibodies among those reporting a prior positive test was 77.3% (58 / 75). The interval between the reported prior positive test date and the antibody test date did not exhibit a strong correlation to the fraction of testing antibody positive, other than perhaps the 3 individuals reporting a very recent (less than 1 week) positive test all testing negative for antibodies (Supplementary Table S2). The rate of survey participants agreeing to antibody testing was lower for those who reported a prior positivetest compared to those who did not: 63.0% (75 of 119) vs. 72.8% (13,923 of 19,128), a small but statistically significant (P < 0.01) difference in proportion.

Of the retained households, 193 (2.1%) had at least one household member who either tested positive for antibodies or reported a prior positive test. There were 159 households with exactly 1 positive member (by either antibody test or reported prior test or both), 26 households with 2 positives, 6 with 3 positives, 1 with 4 positives, and 1 with 6 positives. In all, there were *C* = 273 unique **y**_*i*_ vectors representing household data described in section 2.1.

The crude secondary attack rate measure derived from antibody testing only (fraction of antibody-tested housemates of antibody-positive household members who were also antibody positive) was 14.9% (29 / 194). The crude secondary attack rate estimate from reported prior test data only (fraction of surveyed housemates of people reporting a prior positive test who also reported a prior positive test) was 23.0% (31 / 135). When combining both types of data, the crude secondary attack rate estimate (fraction of surveyed / tested housemates of any antibody-positive or reported-prior-positive person who were positive by either or both measures) was 15.6% (46 / 295).

We tallied demographic statistics of the set of surveyed individuals (Supplementary Table S3). The distribution of reported ages skewed older than Utah’s overall population age distribution, and females were slightly overrepresented (52.0%). The distribution of surveyed individuals’ race, Hispanic origin, and education level also differed from the overall Utah and U.S. distributions.

### 3.2 Maximum likelihood estimates

Our MLE procedure produced simultaneous estimates for all 7 parameters (Table 1). The MLE for *p*_*c*_, the per-person community acquisition probability from outside the household, was 0.41% (0.32% – 0.51%). For within household transmission probability, the MLE produced an average secondary attack rate estimate *p*_*h*_ = 36% (27% – 48%). The MLE for the dispersion parameter *d*_*h*_, quantifying variability in transmissibility by person, was 0.43 (0.02 – 2.0). The boundary case *d*_*h*_ = ∞, representing the classic binomial household transmission model with no variability in individual infectiousness [27], could be rejected with P = 0.001 (Table 2).

**Table 1.** Maximum likelihood estimates

| Value | MLE (95% CI) | Parametric bootstrap:<br>median (95% range) |
| --- | --- | --- |
| Mean community acquisition probability ($p_c$) | 0.41% (0.32% – 0.51%) | 0.41% (0.30% – 0.55%) |
| Mean per-capita household transmission probability ($p_h$) | 36% (27% – 48%) | 36% (25% – 51%) |
| Per-capita household transmission dispersion ($d_h$) | 0.43 (0.02 – 2.0) | 0.38 (0 – 2.2) |
| Probability infected person reported a prior positive test ($\phi_V$) | 72% (62% – 82%) | 72% (63% – 82%) |
| Probability infected person tested positive for antibodies ($\phi_A$) | 86% (75% – 93%) | 86% (77% – 95%) |
| Probability uninfected person did not report a prior positive test ($\pi_V$) | 99.94% (99.88% – 99.98%) | 99.94% (99.87% – 99.99%) |
| Probability uninfected person tested negative for antibodies ($\pi_A$) | 99.3% (99.2% – 99.5%) | 99.3% (99.2% – 99.5%) |

Confidence intervals for MLE derived from the likelihood ratio test, varying each individual parameter while fixing other parameters at their MLE values. Parametric bootstrap was based on MLE fits to 500 different synthetic data sets generated from stochastic simulations using the MLE parameter values.

**Table 2.** Comparison of MLE for alternate models

| Fixed values | $\hat{p}_c$ | $\hat{p}_h$ | $\hat{d}_h$ | $\hat{\phi}_V$ | $\hat{\phi}_A$ | $\hat{\pi}_V$ | $\hat{\pi}_A$ | Log<br>likelihood | Rejection<br>P value |
| --- | --- | --- | --- | --- | --- | --- | --- | --- | --- |
| None | 0.408% | 36.3% | 0.434 | 72.4% | 85.6% | 99.94% | 99.3% | -1173.84 | - |
| $d_h = 0$ | 0.443% | 41.3% | **0\*** | 68.4% | 80.3% | 99.95% | 99.3% | -1174.85 | 0.16 |
| $d_h = \infty$ | 0.340% | 31.9% | $\infty^*$ | 75.6% | 89.0% | 99.91% | 99.3% | -1179.30 | 0.00096 |
| $\pi_V = 100\%$ | 0.508% | 33.7% | 0.161 | 69.9% | 76.9% | **100%\*** | 99.3% | -1176.01 | 0.037 |
| $\pi_A = 99.6\%$ | 0.569% | 30.7% | 0.208 | 59.9% | 82.7% | 99.96% | **99.6%\*** | -1181.08 | 0.00014 |
| $\pi_A = 100\%$ | 1.24% | 17.6% | 0.0217 | 36.6% | 78.4% | 100% | **100%\*** | -1201.26 | <0.0001 |

\*Values that were fixed for the model in that row; other values were optimized by MLE. P values were derived from the likelihood ratio test, compared to the likelihood of the overall MLE in the top row (twice the difference in log likelihood compared to the chi-squared distribution with one degree of freedom).

Our MLE result for *ϕ*_*V*_, the probability that a surveyed person with a prior infection reported a prior positive test, was 72% (62% – 82%). The *ϕ*_*V*_ value can be interpreted as the case ascertainment fraction, i.e. fraction of individuals with SARS-CoV infections who were identified with a positive test during their infection. Our result may be high compared to other areas of the U.S.: one study estimated that less than 60% of symptomatic cases in the U.S. were identified during February-June 2020 [31]. Our finding may reflect unusually successful case ascertainment efforts in Utah during the Spring and early Summer of 2020, perhaps partly owing to slower emergence compared to other regions.

For *π*_*V*_, the probability that a surveyed person with no prior infection reported no prior positive test, the MLE was 99.94% (99.88% – 99.98%). This result is consistent with the low probability of false positives among viral tests, which to our knowledge were exclusively PCR-based in Utah prior to our data collection. It is possible that some false positives in our survey data occurred by erroneous reporting, i.e. survey respondents reporting a prior positive test that did not occur, rather than via errors in testing procedure. Even though our MLE for this parameter was in excess of 99.9%, we found that an alternate model assuming *π*_*V*_ = 100% produced notably different estimates of some of the other parameters (Table 2), which suggests that studies producing epidemiological estimates relying on a 100% viral test specificity assumption should test robustness of conclusions to small deviations from that assumption. For *ϕ*_*A*_, the probability that a prior-infected person’s antibody test was positive, the MLE was 86% (75% – 93%), a result that is similar to the test manufacturer’s finding that 109 of 122 (89%) PCR-positive subjects were positive for antibodies [32]. However, the manufacturer’s results included only symptomatic subjects and were highly dependent on the number of days post-symptom onset at which the serological sample was taken. Because the symptom histories of the antibody-tested people in our data are largely uncertain, it is difficult to determine how consistent our result is with the manufacturer’s data.

For *π*_*A*_, the probability that an antibody-tested person with no prior infection tested negative for antibodies was 99.3% (99.2% – 99.5%), which is within the uncertainty range of the test manufacturer’s estimate of 99.6% (99.0% – 99.9%) based on 4 positive tests from 997 samples collected prior to September 2019 [32]. When instead assuming the manufacturer’s point specificity estimate of 99.6% directly, our estimates of the other parameters changed modestly (Table 2). When we ran our MLE under the assumption of perfect specificity (no false positives) for the antibody test (*π*_*A*_ = 100%), the result for secondary attack rate reduced from 36% to 18%, which is closer to the crude estimate described in Section 3.1, and the results for community acquisition probability *increased* from 0.4% to 1.2% (Table 2). Thus, our model suggests that allowing for false positives can shift the attribution of infections toward household transmissions and away from acquisitions outside the household. We also found that assuming perfect specificity of the antibody test dramatically reduced the estimate of *ϕ*_*V*_ from 72% to 37% (Table 2), which suggests that ignoring false positives in serology data could cause an underestimate of the case ascertainment rate if the serology data are used for that purpose.

When optimizing the likelihood equation against 500 synthetic data sets simulated using the MLE variable assumptions, the median estimates of each parameter were very close to the MLE values (Table 1). The confidence intervals derived from these bootstrap estimates were similar to those derived from the likelihood ratio test, though the bootstrap intervals were somewhat wider for the three parameters governing importation and transmission. Likewise, the likelihood ratio-based intervals reported in Table 1 expanded modestly when we calculated 2-dimensional confidence regions based on each pair of estimated parameters, with most regions exhibiting close to symmetric shapes around the MLE (Supplemental Figures). Notably, the 95% confidence regions involving the transmission dispersion parameter *d*_*h*_ can extend to the high-variability boundary *d*_*h*_ = 0, a result that is also reflected by the fact that the MLE for the model with fixed *d*_*h*_ = 0 cannot be rejected with high confidence (P = 0.16) (Table 2).

Our alternate model that employed a beta-binomial distribution for the number of household acquisitions, using a new dispersion parameter *d*_*c*_ estimated as an additional variable in the MLE, found *d*_*c*_ = 2.1 (0.89 – 7.5), with somewhat altered estimates of the other parameters (Table S4) compared to those in Table 1. However, the log likelihood of the model in Table 1, which is equivalent to the alternate model with *d*_*c*_ = ∞, is sufficiently close to that of the alternate model that *d*_*c*_ = ∞ cannot be rejected by the likelihood ratio test and is favored by the Bayesian information criterion. However, if overdispersion in household community acquisitions does occur, the uncertainty ranges of the transmission variables *p*_*h*_ and *d*_*h*_ become large (see Supplementary Results).

### 3.3 Household transmission variability

We quantified the implications of our key finding of high transmission variability within households of persons infected with COVID-19 by calculating the probability of transmission extremes. Compared to our overall MLE, the classic binomial transmission model (*d*_*h*_ = ∞) produced a similar average secondary attack rate estimate of *p*_*h*_ = 32% (24% – 41%). However, the binomial model produces substantially lower probabilities that an infected individual transmits to no one or everyone in larger households (Table 3).

**Table 3.** Effect of transmission overdispersion on probability that first infected person transmits to no one or everyone in the household

| Household size | Transmit to none: overall<br>model MLE (95% CI) | Transmit to none: binomial<br>model MLE (95% CI) | Transmit to all: overall<br>model MLE (95% CI) | Transmit to all: binomial<br>model MLE (95% CI) |
| --- | --- | --- | --- | --- |
| 2 | 64% (49% – 75%) | 68% (59% – 76%) | 36% (25% – 51%) | 32% (24% – 41%) |
| 3 | 57% (42% – 71%) | 46% (35% – 57%) | 29% (14% – 49%) | 10% (6% – 17%) |
| 4 | 53% (34% – 69%) | 32% (21% – 43%) | 26% (9% – 49%) | 3% (1% – 7%) |
| 5 | 51% (30% – 69%) | 22% (12% – 33%) | 24% (6% – 50%) | 1% (0.3% – 3%) |
| 6 | 49% (26% – 69%) | 15% (7% – 25%) | 22% (5% – 50%) | 0.3% (0.08% – 1%) |
| 7 | 47% (24% – 69%) | 10% (4% – 19%) | 21% (3% – 50%) | 0.1% (0.02% – 0.5%) |
| 8 | 46% (22% – 70%) | 7% (3% – 14%) | 20% (3% – 50%) | 0.03% (0.005% – 0.2%) |
| 9 | 45% (20% – 70%) | 5% (2% – 11%) | 20% (2% – 50%) | 0.01% (0.001% – 0.08%) |
| 10 | 44% (19% – 70%) | 3% (0.9% – 8%) | 19% (2% – 51%) | 0.003% (0.0003% – 0.03%) |

Probabilities in this table are for a single infected household member transmitting directly to no one or everyone else in the household. The
“transmit to all” values do not include the probability of multiple-generation transmission chains that eventually infect all household members.
Confidence intervals for the overall MLE-based estimates were derived from applying $(p_h, d_h)$ pairs from our parametric bootstrap analysis to the beta-binomial transmission equations.

For example, our MLE model estimates that an infected member of an 8-member household would have a 46% (22% – 70%) chance of transmitting to no one, but a 20% (3% – 50%) chance of transmitting infection directly to all 7 housemates. By contrast, the no-variability binomial model estimate would be substantially lower for each extreme: 7% (3% – 14%) chance of transmitting to no one and 0.03% (0.005% – 0.2%) chance of transmitting to everyone (Table 3).

We calculated an example of a dynamic transmission model that would produce the same mean and variance of a person’s transmission probability to a household member that is produced by our MLE beta distribution. If an infected person’s duration of infectiousness is assumed to be fixed and transmissibility to a housemate is modeled as a gamma distribution with shape *k*, then *k* = 0.18 (95% CI 0 – 0.7) when the mean and variance are matched, regardless of the infectious duration (Supplementary Methods). This estimate of *k* is comparable to the dispersion parameter *k* of the negative binomial distribution commonly used to characterize overall variability in the number of transmissions from individuals, which can be derived from the Poisson distribution with a mean that is gamma-distributed with shape parameter *k* [17]. Our estimate of *k* is similar to point estimates for SARS-CoV-2 of *k* = 0.1 [22], *k* = 0.25 [23], and *k* = 0.33 [24].

### 3.4 Within-household reproduction numbers

Our estimate of the household reproduction number *R*_*h*_, the expected number of household transmissions from a community acquirer with no other infected fellow household members, depends on our estimate of *p*_*h*_ and the mean *μ* and variance *σ*^2^ of the household size distribution. From our data we found *μ* = 3.07 and *σ*^2^ = 3.12, so our estimate is *R*_*h*_ = 1.12 (0.78 – 1.56). Our estimate of the alternate household reproduction number 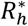, the expected total number of transmissions in the household of a community acquirer, is 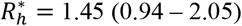.

The supercritical threshold for *R*_*c*_, the average number of non-household transmissions by an infected individual, is approximated by 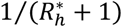 (see Methods section 2.6 and Supplementary Material). Using our estimate for 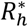 in Utah, this formula suggests that *R*_*c*_ must be kept below approximately 0.41 (0.33 – 0.52) to avoid increasing growth of COVID-19 infections in the population.

## 4. Discussion

The key findings of our analyses stem from our simultaneous estimation of the average and variability of SARS-CoV-2 household transmission, household importation, and test data accuracy. Our novel combination of those interacting features within our model revealed two important epidemiological insights. First, we found that accounting for test error, especially the specificity of the serological antibody test, produced a substantially higher estimate for the household secondary attack rate. Second, we found evidence of substantial variability of transmissibility within households, which has important implications for understanding broad transmission patterns and mitigation strategies.

An important implication of the first finding is that assuming perfect test accuracy may be a source of underestimation for the household secondary attack rate in other studies. Our maximum likelihood estimate was 35% (27% – 48%), which is higher than recent pooled estimates of 17–19% from the most recent meta-analyses of worldwide household studies [7, 8]. These and other published studies have generally estimated the secondary attack rate by a simple calculation of the fraction of tests that were positive among household contacts of known cases. When we applied that calculation to our combined data, we found a crude secondary attack rate estimate of 15.6%. We traced the major source of this substantial underestimate to the assumption of perfect test specificity inherent in the crude formula.

[32]Our second major finding of overdispersion of household transmission stemmed from our use of the beta-binomial distribution to quantify the number of household transmissions from infected individuals. We quantified individual-level variability in transmissibility using a dispersion parameter *d*_*h*_, and the optimal value occurred at low dispersion (high variability; *d*_*h*_ = 0.43). The more commonly used binomial model, a special case of our model at minimal variability (*d*_*h*_ → ∞), was rejected, suggesting that transmission patterns are not well captured by that simplifying assumption.

Our dispersion parameter estimate is not directly comparable to another commonly used dispersion parameter, often named *k*, that characterizes variability in the total number of transmissions (whether household or not) from each infected person as a parameter of the negative binomial distribution [17]. We converted *d*_*h*_ to *k* in the context of simple model in which the only source of variability is a person’s transmissibility per unit time in contact with others, finding *k* = 0.18, similar to other published results for SARS-CoV-2. This similarity perhaps suggests that variability in infectivity per time is a major driver of overall transmission variability for SARS-CoV-2. This could be consistent with findings that viral shedding is highly variable by individuals with SARS-CoV-2 infections, both during asymptomatic and symptomatic phases of disease, suggesting that heterogeneous transmissibility may be largely explained by overdispersion in levels of viral shedding by individuals [33]. However, other studies suggest that SARS-CoV-2 transmission overdispersion in the wider population beyond households may be less driven by biological heterogeneity and more by heterogeneous social contact behavior [34].

The level of within-household transmission variability captured by the parameter *d*_*h*_ affects the contribution of household transmission toward threshold levels of overall transmission. Threshold conditions are often expressed using a reproduction number (*R*), the average number of transmissions from each infected person. The average number of household transmissions directly from an initially infected household member (*R*_*h*_) is independent of *d*_*h*_, but *d*_*h*_ does affect the average number of household transmissions in the next generation, i.e. by someone who acquired infection from a housemate. When transmission variability is higher, the household transmission potential of a household acquirer is lower, reducing to zero in the “all-or-nothing” limit *d*_*h*_ = 0. To capture this effect, we introduced an alternate reproduction number 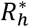, which is the average number of total household transmissions after the initial introduction, when final household outbreak size has been reached.

Neither *R*_*h*_ > 1 nor 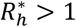 are sufficient threshold conditions for sustained transmission in a community, which requires some level of between-household transmission to be maintained. Given our estimate of 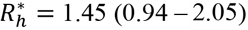, we can estimate the critical value of *R*_*c*_, the average number of non-household community transmission that would push transmission for the population above the supercritical threshold for a growing epidemic, with the threshold condition 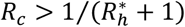. Thus, we estimate that *R*_*c*_ must be kept below approximately 0.41 (0.33 – 0.52) to avoid continued case growth in Utah if household transmission continues to be well characterized by our model. As this result depended on the average household size in our data, it is notable that Utah has the highest state-average household size in the United States. The average household size in Utah is 3.1, about 20% higher than the national average household size. Thus, our *R*_*h*_ estimate may be high compared to other locations. A lower value of *R*_*h*_ would lead to a higher threshold value for *R*_*c*_.The potential contribution of interventions to reduce household transmission may also be important. Using the terms defined above, if *R*_*c*_ < 1 but 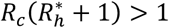, then overall transmission is above-threshold but could be pushed below-threshold by reducing household transmission alone, such that 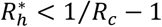. Methods to reduce household transmission might include increased used of at-home testing to earlier detect potential asymptomatic or pre-symptomatic transmitters, paired with increased use of masks, disinfectants, and/or distancing within homes of an infectious person [35]. [32][31]

This study has several limitations. Our estimate of high household transmission variability may not be robust to alternate assumptions for the way community acquisition risk varies by household. For example, some households could have been comprised of families with both parents working essential jobs during Spring/Summer 2020, with children attending in-person day care or camps, thus placing the entire household at much higher risk of community acquisition compared to households working / caring for children at home. Also, households could have high collective community acquisition probability via attending multi-household gatherings of extended family or other social groups. In these ways, households conceivably could vary considerably in their infection numbers for reasons that don’t involve within-household transmission.

We tested the implications of this alternate possibility for household variability in our model by allowing variability in community acquisition by household using an additional dispersion parameter to the MLE model (Supplementary Results). Interestingly, the MLE for the transmission dispersion parameter *d*_*h*_ still occurred at high variability in household transmission 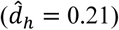 under this alternate model. Furthermore, the improvement in likelihood was not substantial, such that the more complicated model would not be favored by the likelihood ratio test nor the Bayesian information criterion. However, larger uncertainty ranges under the alternate model suggest that we may not be able to definitively rule out the possibility that variability in community acquisition risk by household plays a substantial role in explaining overall variability in household infection numbers.

It is also possible that household transmission variability could be driven by properties of households such as contact behavior, underlying health composition of household members, physical properties of the domicile such as size and ventilation, or other properties that could increase transmission risk of all household members together. Possible variability in person-to-person transmission probability by household, rather than by individual, is not accounted for in our model. Using a beta distribution for this probability across different households to arrive at an alternate final size distribution would require integrating the beta distribution over the full final size distribution equations produced by the binomial-chain model, which would be complicated for larger households. Alternatively, one could model a functional relationship between observed properties of a household in the dataset and its average transmission probability, while retaining dispersion occurring at the individual level. We have not attempted this with our data; we suspect that the sample size of outbreaks in households with a given feature would not be large enough to draw meaningful conclusions, but this could be an important direction of future work enhanced by a larger dataset.

Another limitation lies in our potentially inaccurate assumptions used to quantify the probability of prior infections among those with missing data within participating households. Most non-participating individuals within participating households were children under 12, who were not offered antibody tests. Older participants could fill out surveys on behalf of children of any age, including reporting of prior positive tests, but participation in that option was low. Thus, our assumption that non-participants had equal community acquisition rates, susceptibility to acquisition from another household member, and transmissibility to other household members compared to study participants would be violated if children were substantially different from adults in one or more of those quantities. Our assumption is consistent with studies finding similar transmission rates to and from children compared to adults. In a study of COVID-19 clusters linked to day care centers within our study area in Utah [36], 42% of the cases occurred in children, who represented 60% of the people with epidemiological contacts to the facilities. The infected children (median age 7) transmitted infection to at least 26% of their non-facility contacts, close to our household estimate. Another study found that children under 10 in China were as likely to be infected as adults [37]. However, other studies suggest that children may be less likely to acquire infection than adults [38], and one study found very low household secondary attack from infected children in South Korea [39]. A study similar to ours found lower rates of importation and household acquisition among children aged 5-9 compared to older groups, although confidence intervals overlapped [15]. If substantial differences existed between children under 12 and our study participants, one or more of our estimates could be biased.

In addition, many eligible participants older than 12 chose not to participate, either declining the serological antibody test only (but still filling out a survey) or declining to participate at all. Comparing full participants to survey-only participants, we found that participants reporting a prior positive SARS-CoV-2 test were less likely to agree to antibody testing, though the difference was not large (63.0% vs. 72.8%). It is unknown whether a prior confirmed or suspected infection affected eligible household members’ decision to agree or decline to fill out a survey. The full set of surveyed participants had different distributions of reported age, sex, race, Hispanic origin, and education level compared to the wider population, and future work could assess the implications of those differences for extrapolating COVID-19 risk to other households.

We also have not adjusted for potential biases related to non-participation rates of entire households that were selected and approached for inclusion in the study. Our data collection included a complicated sampling design across several different strata, and weights were introduced partly to account for different rates of nonresponse across the different strata. For simplicity we ignored these details and sampling weights for the analysis presented here. Also, while the 7 included Utah counties represent >86% of the state population, there may be important differences in households from the 22 excluded counties. Thus, households with higher COVID-19 risk may be overrepresented or underrepresented in our data relative to their frequency in the broader population of households in Utah.

Although these potential limitations, which also exist for other analyses of household transmission from serological data [13-15], remain in our analysis, we believe our model has addressed other limitations of existing models that may be more substantial. Our improvements to household secondary attack rate estimates, including factoring out non-household community acquisitions and tertiary transmissions, inclusion of overdispersion estimates, and careful consideration of the impact of imperfect test sensitivity and specificity, have produced improved insights into this important measure. While the likelihood equations resulting from our model are somewhat complicated, we have provided full mathematical specification and computational code for reproducibility. The ability to explicitly calculate the likelihood for our model is an advantage for optimization speed and further mathematical analysis, and extensions to the epidemiological household model can readily be simulated to explore potential improvements.

In conclusion, we found evidence of a relatively high secondary attack rate and high overdispersion in transmission of SARS-COV-2 in Utah households during a time when overall community prevalence was low. Other published household secondary attack rates may be underestimated without accounting for imperfect test sensitivity and specificity. Controllability of the virus may depend on mitigating transmission from a minority of highly infectious individuals in large households and other household-like locations where several people congregate indoors for extended periods.

## Data Availability

Data necessary to reproduce the results in the manuscript are publicly available.

https://github.com/damontoth/householdTransmission

## Supplementary material

### Supplemental Methods

#### Survey data

Answers to the following questions for individual household members were used in our analysis:

1. “Have you ever been tested for coronavirus (also called SARS-CoV-2 or COVID-19)?” (Yes or No) If the answer to 1) was Yes, the following two questions were asked:
2. What was the result? (Positive; Negative; Have not received test result; or Don’t know)
3. When were you tested? (MM/DD/YYYY)

All individuals who answered “Yes” to question 1 and “Positive” to question 2 were classified as “reported a prior positive test,” and all other surveyed individuals were classified as “did not report a prior positive test,” as described in the main text. For individuals who reported a prior positive test and also received an antibody test, we used the answer to question 3, compared to the collection date of serology, to construct Table S2.

#### Alternate model with variability in household importation

For the alternate model, the formula for *M*_*kn*_ for a given household size *n* ≥ 2 becomes

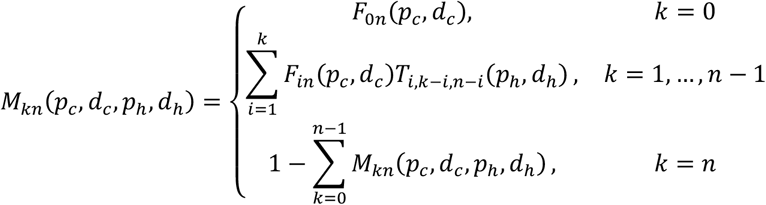

For households of size *n* = 1, *M*_01_(*p*_*c*_, *d*_*c*_) = *F*_01_(*p*_*c*_, *d*_*c*_) and *M*_11_(*p*_*c*_, *d*_*c*_,) = 1 − *F*_01_(*p*_*c*_, *d*_*c*_). The function *F*_*yz*_(*p, d*) is defined in the main text (probability mass function of the beta-binomial distribution with boundary case definitions at *d* = 0 and *d* → ∞), where in this case *y* is the number of community acquisitions and *z* is the total number of household members. The main-text model is a special case of this alternate model, with *d*_*c*_ → ∞.

The likelihood equation is the same as in the main text, but with the additional element *d*_*c*_ in the vector **θ** of variables to be optimized:

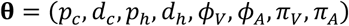

and *M*_*kn*_(*p*_*c*_, *p*_*h*_, *d*_*h*_) in the likelihood equation is replaced with *M*_*kn*_(*p*_*c*_, *d*_*c*_, *p*_*h*_, *d*_*h*_) as defined above.

We found the MLE and single-parameter confidence intervals using the same procedure described in the main text, and further assessed uncertainty of *d*_*h*_ by solving for the MLE of the other 7 variables when fixing it at its boundary values 0 and ∞. We also compared the likelihood at the MLE of the alternate model to that of the main text model using the likelihood ratio test, to determine whether the main text model result could be rejected in favor of the alternate model by this criterion. As an additional comparison, we used the Bayesian information criterion to score the alternate model against the main text model, using 9224 as the number of data points (number of households) and 7 and 8 as the number of parameters for the main-text model and alternate model, respectively.

#### Beta-binomial distribution at limits of dispersion parameter: d → ∞ and d → 0

Our likelihood equations make use of the beta-binomial probability distribution, parameterized with an average probability *p* and a dispersion parameter *d*. The probability mass function *F* for positive, finite values of *d* is

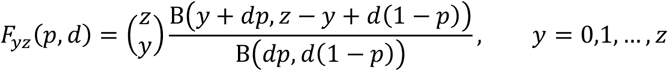

We use *F*_*yz*_(*p, d*) to quantify the distribution of household transmissions directly from a single infected household member, where *y* is the number of transmissions, *z* is the number of susceptible household members, *p* = *p*_*h*_, and *d* = *d*_*h*_. In our alternate model we also use *F*_*yz*_(*p, d*) to quantify the distribution of community acquisitions among members of a household from non-household members, where *y* is the number of community acquisitions, *z* is the total number of household members, *p* = *p*_*c*_, and *d* = *d*_*c*_.

Here, we derive the formula for *F*_*yz*_(*p, d*) at the boundaries of the range of possible values for *d*: *d* → ∞ and *d* → 0. To do this, we rewrite *F*_*yz*_(*p, d*) in an alternate form. First, using the property B(*x, y*) = Γ(*x*)Γ(*y*)/Γ(*x* + *y*):

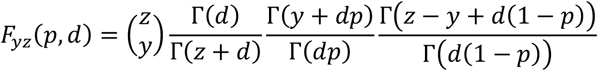

Then using the property, for positive integer *n*, Γ(*z* + *n*) = *z*(*z* + 1) … (*z* + *n* − 1)Γ(*z*):

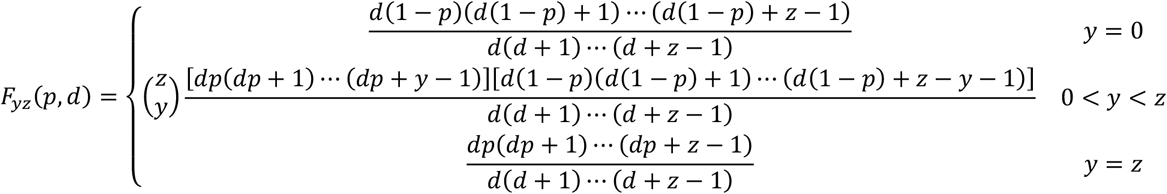

In the following we rewrite the numerators and denominators in powers of *d*. Only the lowest and highest powers of *d* will matter for taking the limits that follow, so the other terms within “…” are not shown.

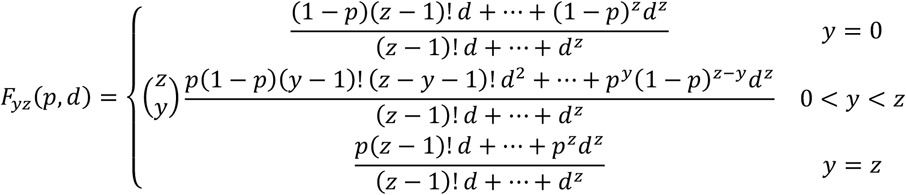

When taking the limit *d* → ∞, we note that each fraction has highest order *d*^*z*^ in both numerator and denominator, so the limit will be the ratio of the coefficients of that term:

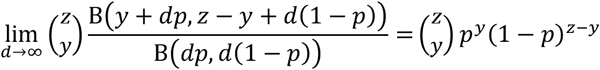

When taking the limit *d* → 0, we note that the fraction for cases *y* = 0 and *y* = *z* have lowest order term *d* in both the numerator and denominator, so the limit will be the ratio of the coefficients on those terms. The fraction for the 0 < *y* < *z* case has only powers of *d*^2^ and higher in the numerator, and a nonzero *d* term in the denominator, so the limit is 0:

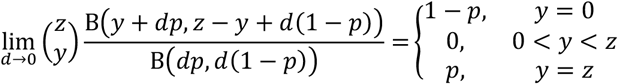

#### Direct transmission probabilities H_xyz_

Here we derive the formulae for *H*_*xyz*_: the probability of *y* transmissions to *z* susceptible household members directly from *x* infected members

As described in the main text, we first define the probabilities for transmissions directly from *x* = 1 infected household member:

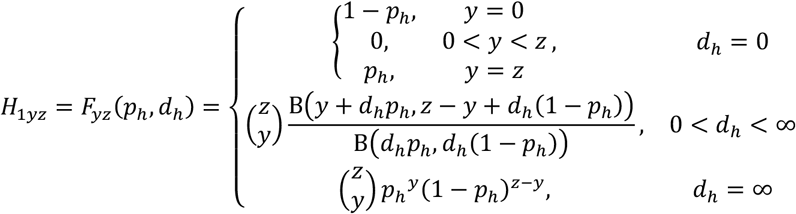

Next consider *x* = 2 infected household members. The probability that *y* = 0 transmissions occur is the probability that both infected members transmit to 0 others: *H*_20*z*_ = *H*_10*z*_*H*_10*z*_. To calculate the probability that *y* > 0 transmissions occur from the two infected members, it is convenient to consider the two infected individuals having transmission opportunities in sequence, say A followed by B. If A transmits to any household members, this reduces the number of susceptible members remaining for B to infect. For example, the probability that *y* = 1 is the probability that A transmits to 0 of *z* and B transmits to 1 of *z*, plus the probability that A transmits to 1 of *z* and B transmits to 0 of *z* − 1 remaining susceptible members: *H*_21*z*_ = *H*_10*z*_*H*_11*z*_ + *H*_11*z*_*H*_1,0,*z*−1_. The calculation follows a similar pattern for *y* = 2: *H*_22*z*_ = *H*_10*z*_*H*_12*z*_ + *H*_11*z*_*H*_1,1,*z*−1_ + *H*_12*z*_*H*_1,0,*z*−2_. It follows that:

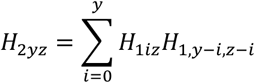

Now for *x* = 3 infected household members, we can use the fact that we have already calculated *H*_2*yz*_, which covers the transmission probabilities from two of the three infected members, and then we include the probability that third member transmits to any remaining susceptible members that the first two did not infect:

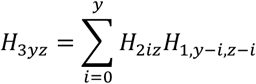

Following this pattern, we continue calculating *H*_*xyz*_ for each *x* in increasing sequence:

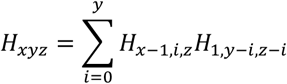

#### Total transmission probabilities T_xyz_

Here we derive the formulae for *T*_*xyz*_: the probability of *y* total transmissions to *z* initially susceptible members from *x* initially infected members. In other words, *T*_*xyz*_ is the probability that the final household outbreak size is *x* + *y*, given that *x* household members were initially infected and *z* household members were initially susceptible.

First, we note that *T*_*x*0*z*_ = *H*_*x*0*z*_ for all possible (*x, z*) pairs, because if the initial *x* infected members do not transmit to anyone (*y* = 0), the household outbreak is over and the final size has been reached. Next we consider the probability of *y* = 1 total transmissions, which occurs when the initial *x* infected members transmit directly to 1 other (with probability *H*_*x*1*z*_), who then does not subsequently transmit to any of the remaining *z* − 1 susceptible members (with probability *H*_1,0,*z*−1_ = *T*_1,0,*z*−1_). Hence,

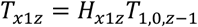

For *y* = 2 total transmissions, we must include the probability that the initial *x* infected members transmit directly to 2 others who then transmit to none and the probability that the initial *x* infected members transmit directly to 1 other who then produces an outbreak among the remaining susceptible members with 1 total transmission:

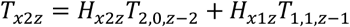

Following similar logic for *y* = 3, and making use of the *T*_*x*0*z*_, *T*_*x*1*z*_, and *T*_*x*2*z*_ values already calculated, we arrive at:

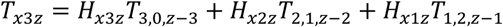

The general formula calculated for increasing values of *y* is:

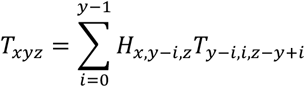

#### Within-household reproduction numbers and threshold condition

We define the within-household reproduction number *R*_*h*_ as the expected number of household transmissions directly from an infected person who acquired infection in the community and has no non-susceptible housemates. Let *h*_*i*_ be the fraction of households with size *i*, up to a maximum size *N*. Then the mean *μ* and variance *σ*^2^ of the household size distribution are

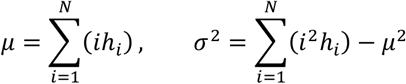

Let *c*_*i*_ be the probability that a randomly chosen person has *i* housemates. The probability that a randomly chosen person lives in a house of *total* size *i* (including themselves) is *ih*_*i*_/*μ*, so

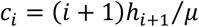

Then *R*_*h*_ is *p*_*h*_ times the mean number of housemates of a randomly chosen person:

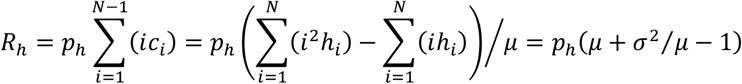

We also define an alternate within-household reproduction number 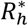 as the expected total number of transmissions in the household of an infected person who acquired infection in the community and has no initially non-susceptible housemates. Given that a person acquiring infection in the community has *i* susceptible housemates, the probability that *j* of their housemates become infected before the household outbreak terminates is *T*_1*ji*_(*p*_*h*_, *d*_*h*_), as defined in Section 2.2 of the main text. The expected total number of transmissions in their household will be then be 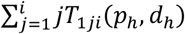. So, the household reproduction number formula is:

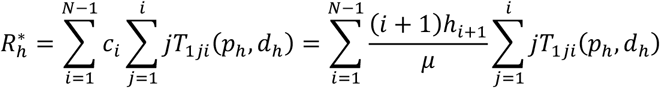

For the high-variability boundary case at *d*_*h*_ = 0, we have that

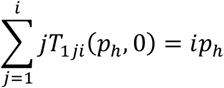

because *T*_1*ji*_(*p*_*h*_, 0) = *p*_*h*_ when *j* = *i* and 0 for other nonzero values of *j* (reflecting all-or-nothing transmission). It follows that 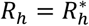 when *d*_*h*_ = 0. This makes intuitive sense because in the all-or-nothing scenario, when the index person transmits, all in the household are infected directly, and there is no one left to infect in subsequent generations, so the final household outbreak size is entirely reflected in *R*_*h*_.

We next investigate the implications of our 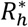 estimate for population-wide transmission control. The threshold condition delineating subcritical and supercritical transmission in the population occurs when the maximal eigenvalue of the matrix

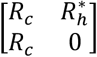

exceeds one [30]. Here, *R*_*c*_ is defined as the average number of community transmissions per infected individual (i.e., average number of transmissions to people not in the infected individual’s household). The occurrence of *R*_*c*_ in both rows of the matrix reflects an assumption that its value applies to the transmissibility of people who acquire their own infection in the community and in their household. The zero element in the lower-right corner of the matrix reflects the fact that the 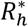 people on average who acquire infection in their household do not transmit further in their household, by definition, because 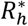 was derived from the final household outbreak size equations encompassed in *T*_*xyz*_.

The maximal eigenvalue exceeding one produces the following threshold condition:

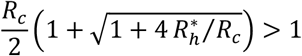

This is equivalent to

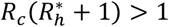

If the threshold condition is met and 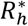 is fixed, then the system can be pushed below threshold by reducing *R*_*c*_ such that

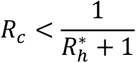

If *R*_*c*_ is fixed and less than one, then the system can be pushed below threshold by reducing 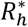 such that

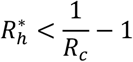

#### Relationship between beta distributed probability and dynamic transmission parameters

If an infected person’s duration of infectiousness is *τ* and the transmission rate to a contact is *β*, the probability that transmission to the contact occurs is *p* = 1 − *e*^−*βτ*^. We assume *τ* is fixed and *β* is a gamma distributed random variable with shape *k* and rate *r*. Then, the first and second moments of the random variable *p* are:

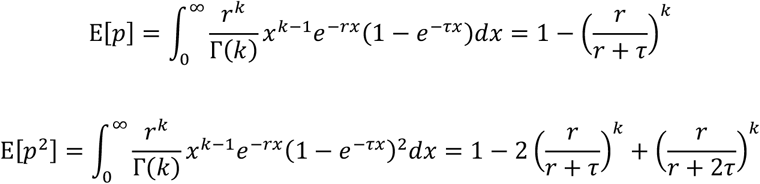

The variance is then

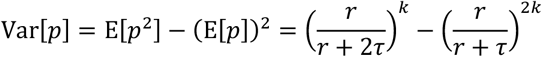

We then equate the mean and variance to those of the beta distribution with mean *p*_*h*_ and dispersion *d*_*h*_, which we used in our MLE model in the main text.

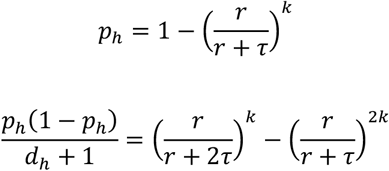

Combining those two equations yields

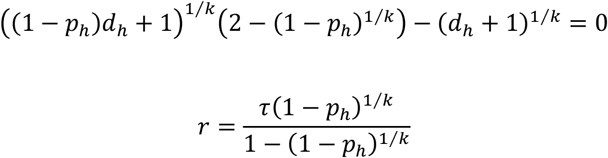

We solved the first equation for *k*, which is independent of the assumption for *τ*, using our MLE estimates of *p*_*h*_ and *d*_*h*_. We applied each of the (*p*_*h*_, *d*_*h*_) pairs from our parametric bootstrap analysis to this equation to derive the confidence interval for *k*.

## Supplemental Results

The alternate model produced an estimate for the new dispersion parameter *d*_*c*_ = 2.1 (0.89 – 7.5) and altered estimates for the other 7 parameters compared to their values for the main text model (Table S4). The log likelihood at this MLE was about 1 greater than the log likelihood produced by the main text result (Table S5), suggesting that the main text model (equivalent to the alternate model with *d*_*c*_ = ∞) cannot be rejected with high confidence in favor of the alternate model by the likelihood ratio test (P = 0.14). The Bayesian information criterion (BIC) for the alternate model is 2418.5 compared to 2411.6 for the main text (*d*_*c*_ = ∞) model, which favors the main text model with a BIC difference of 6.9.

The conclusion of high household transmission variability from the main-text model is consistent under this alternate model, with the MLE occurring at low value of the dispersion parameter *d*_*h*_ = 0.30, and the low-variability binomial model *d*_*h*_ = ∞ can be rejected with P = 0.02. However, uncertainty ranges become wider at higher levels of overdispersion in household risk of community acquisition. This is illustrated by the fact that a model assuming no household transmission (*p*_*h*_ = 0), i.e. all household cases explained by acquisitions outside the households with high overdispersion (*d*_*c*_ = 0.5), cannot be rejected with very high confidence (P = 0.068). Thus, the alternate explanation for the distribution of household cases may not be definitively ruled out by our data.

**Table S1.**
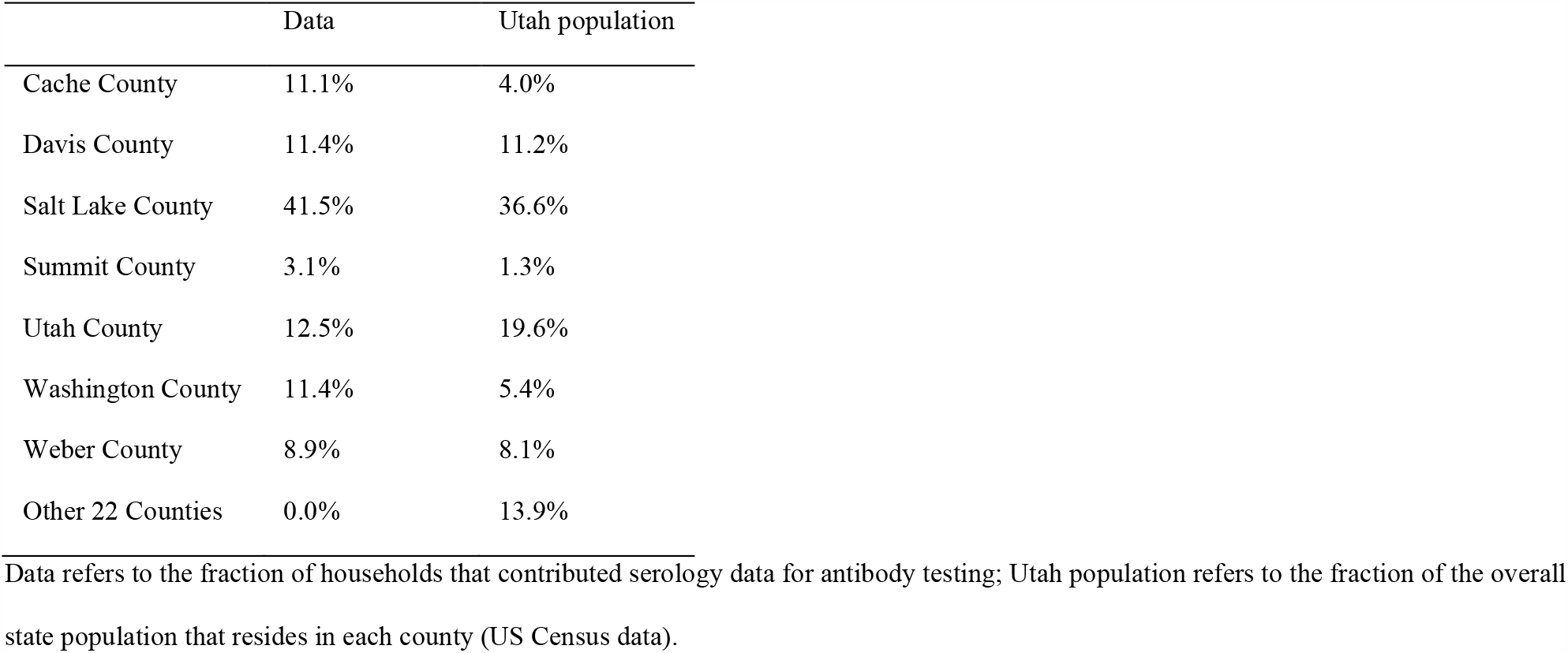
Fraction of household data from each county in the state of Utah

|  | Data | Utah population |
| --- | --- | --- |
| Cache County | 11.1% | 4.0% |
| Davis County | 11.4% | 11.2% |
| Salt Lake County | 41.5% | 36.6% |
| Summit County | 3.1% | 1.3% |
| Utah County | 12.5% | 19.6% |
| Washington County | 11.4% | 5.4% |
| Weber County | 8.9% | 8.1% |
| Other 22 Counties | 0.0% | 13.9% |

Data refers to the fraction of households that contributed serology data for antibody testing; Utah population refers to the fraction of the overall state population that resides in each county (US Census data).

**Table S2.**
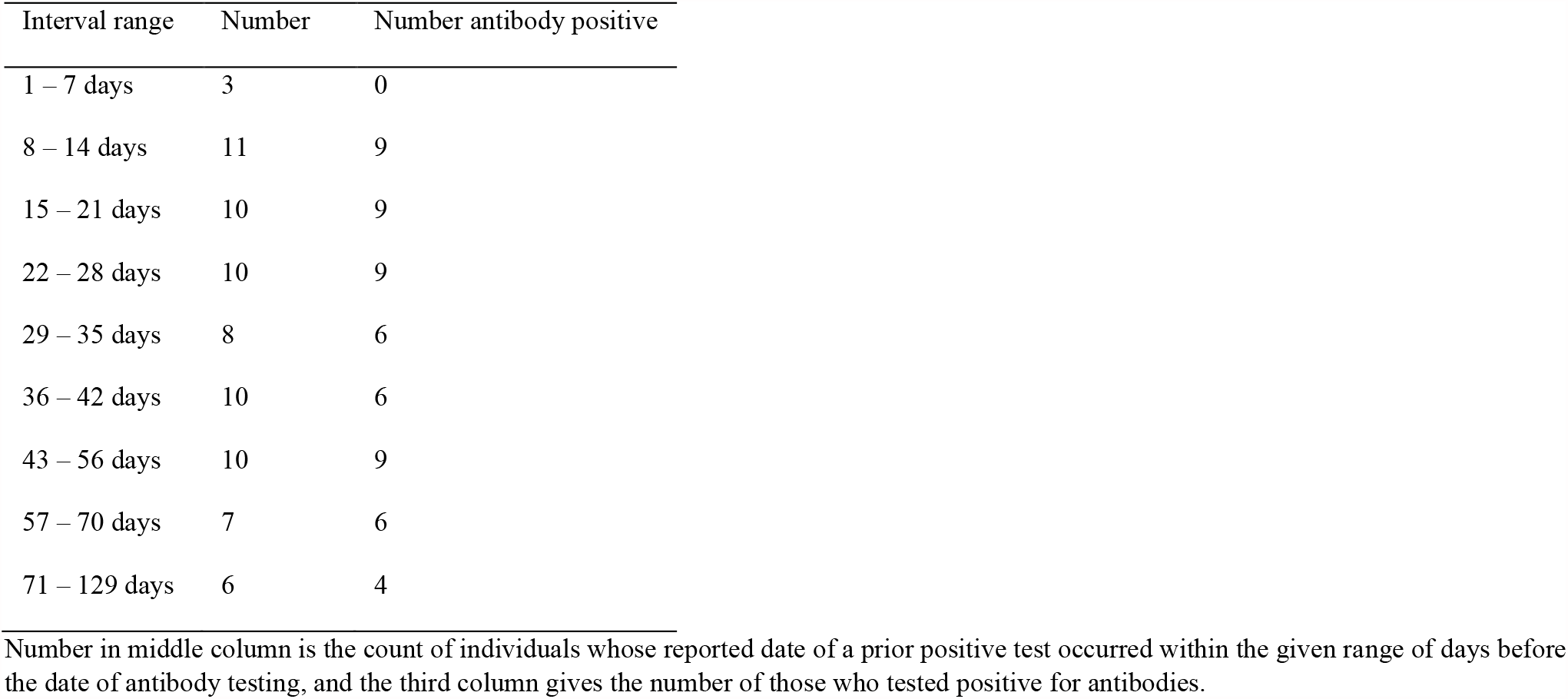
Intervals between reported prior positive test and antibody test results

**Table S3.** Demographic distributions of surveyed individuals

|  | Data | Utah | U.S. |
| --- | --- | --- | --- |
| **Age and Sex** |  |  |  |
| Age 0 to 4 years | 1.6% | 7.7% | 6.1% |
| Age 5 to 14 years | 6.6% | 16.4% | 12.6% |
| Age 15 to 24 years | 13.2% | 16.2% | 13.0% |
| Age 25 to 34 years | 15.8% | 14.7% | 14.0% |
| Age 35 to 44 years | 16.6% | 13.8% | 12.6% |
| Age 45 to 54 years | 12.5% | 10.2% | 12.6% |
| Age 55 to 64 years | 13.8% | 9.5% | 12.8% |
| Age 65 to 74 years | 13.0% | 6.8% | 9.8% |
| Age 75 to 84 years | 5.7% | 3.4% | 4.8% |
| Age 85 years and over | 1.2% | 1.2% | 1.8% |
| Female persons | 52.0% | 49.6% | 50.8% |
| **Race and Hispanic Origin** |  |  |  |
| White alone | 93.8% | 90.6% | 76.3% |
| Black or African American alone | 0.7% | 1.5% | 13.4% |
| American Indian and Alaska Native alone | 0.6% | 1.6% | 1.3% |
| Asian alone | 2.3% | 2.7% | 5.9% |
| Native Hawaiian and Other Pacific Islander alone | 0.5% | 1.1% | 0.2% |
| Two or More Races | 2.0% | 2.6% | 2.8% |
| Hispanic or Latino | 8.5% | 14.4% | 18.5% |
| White alone, not Hispanic or Latino | 87.9% | 77.8% | 60.1% |
| **Education** |  |  |  |
| High school graduate or higher, persons age 25+ | 98.2% | 92.3% | 88.0% |
| Bachelor's degree or higher, persons age 25+ | 57.1% | 34.0% | 32.1% |

Data refers to the fraction of surveyed individuals who reported each characteristic in our survey results. Utah and U.S. columns contain data from the US Census (Vintage 2019 Population Estimates Program).

**Table S4.** Alternate model results: allowing variability in importation probability by household

| Value | MLE estimate (95% interval) |
| --- | --- |
| Mean community acquisition probability ($p_c$) | 0.56% (0.44% – 0.70%) |
| Household community acquisition dispersion ($d_c$) | 2.1 (0.89 – 7.5) |
| Mean per-capita household transmission probability ($p_h$) | 27% (16% – 41%) |
| Per-capita household transmission dispersion ($d_h$) | 0.21 (0 – 3.4) |
| Probability that surveyed, infected person reported a prior positive test ($\phi_V$) | 72% (62% – 82%) |
| Probability that antibody test of person with prior infection was positive ($\phi_A$) | 87% (77% – 94%) |
| Probability that surveyed, uninfected person did not report a prior positive test ($\pi_V$) | 99.92% (99.86% – 99.97%) |
| Probability that antibody test of person with no prior infection was negative ($\pi_A$) | 99.3% (99.2% – 99.5%) |

Confidence intervals for MLE derived from the likelihood ratio test, varying each individual parameter while fixing other parameters at their MLE values.

**Table S5.** Comparison of alternate model MLE to main-text and other models

| Fixed values | $\hat{p}_c$ | $\hat{d}_c$ | $\hat{p}_h$ | $\hat{d}_h$ | $\hat{\phi}_V$ | $\hat{\phi}_A$ | $\hat{\pi}_V$ | $\hat{\pi}_A$ | Log likelihood | # of optimized parameters | Rejection P value |
| --- | --- | --- | --- | --- | --- | --- | --- | --- | --- | --- | --- |
| None | 0.56% | 2.1 | 27% | 0.21 | 72% | 87% | 99.92% | 99.3% | -1172.75 | 8 | - |
| $d_c = \infty$ | 0.41% | $\infty^*$ | 36% | 0.43 | 72% | 86% | 99.94% | 99.3% | -1173.84 | 7 | 0.14 |
| $d_h = 0$ | 0.60% | 1.7 | 24% | **0\*** | 72% | 87% | 99.93% | 99.3% | -1172.86 | 7 | 0.64 |
| $d_h = \infty$ | 0.77% | 0.5 | 0% | $\infty^*$ | 72% | 88% | 99.92% | 99.3% | -1175.29 | 7 | 0.024 |
| $p_h = 0\%$ | 0.77% | 0.5 | **0%\*** | N/A | 72% | 88% | 99.92% | 99.3% | -1175.29 | 6 | 0.079 |

\*Values that were fixed for the model in that row; other relevant values were optimized by MLE. P values were derived from the likelihood ratio test, compared to the likelihood of the overall MLE in the top row: twice the difference in log likelihood compared to the chi-squared distribution with degrees of freedom equal to the difference in the number of optimized parameters. The model with household transmission probability $p_h$ fixed at 0% optimized only 6 parameters because the transmission dispersion parameter $d_h$ is irrelevant with no transmission, hence we used 2 degrees of freedom for the reference chi-squared distribution when comparing the likelihood to the full 8-parameter model. For the model with $d_h$ fixed at ∞ (binomial transmission model), the optimum occurred at the boundary $p_h = 0$ , producing the same likelihood as the adjacent model but different P-value as the reference chi-squared distribution has 1 degree of freedom.

**Figure S1.**
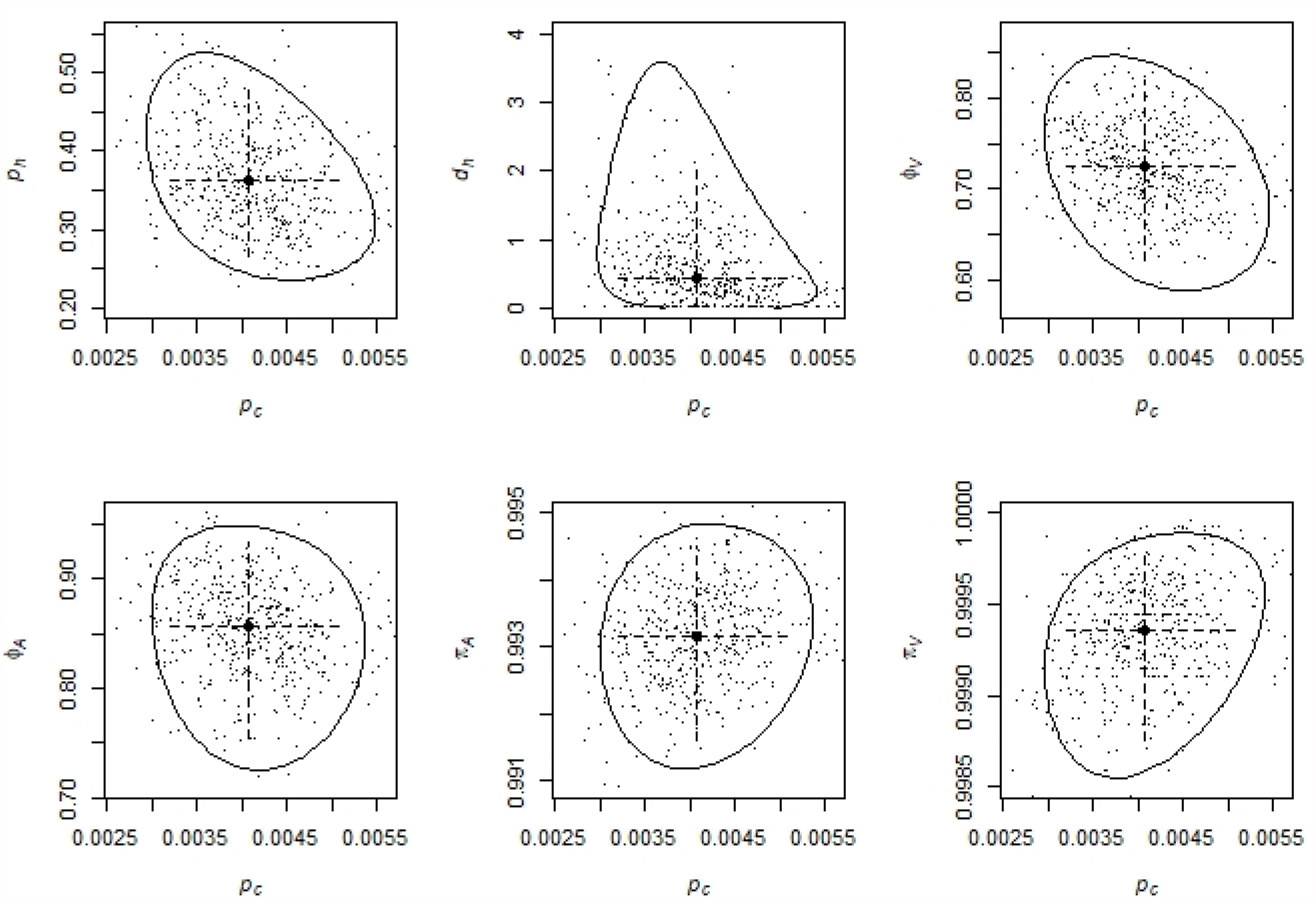
Two-dimension confidence regions for *p*_*c*_ paired with each other parameter Solid curves are the 2-dimensional confidence regions derived from the likelihood ratio test, comparing the likelihood ratio statistic to the 95^th^ percentile of the chi-squared distribution with 2 degrees of freedom. Large solid circle is the MLE estimate and dashed lines are the confidence intervals for each individual parameter derived from the likelihood ratio test (Table 1 main text). Small dots are the MLE estimates from each of 500 simulated data sets generated using parameter values set at the MLE from the actual data (parametric bootstrap).

**Figure S2.**
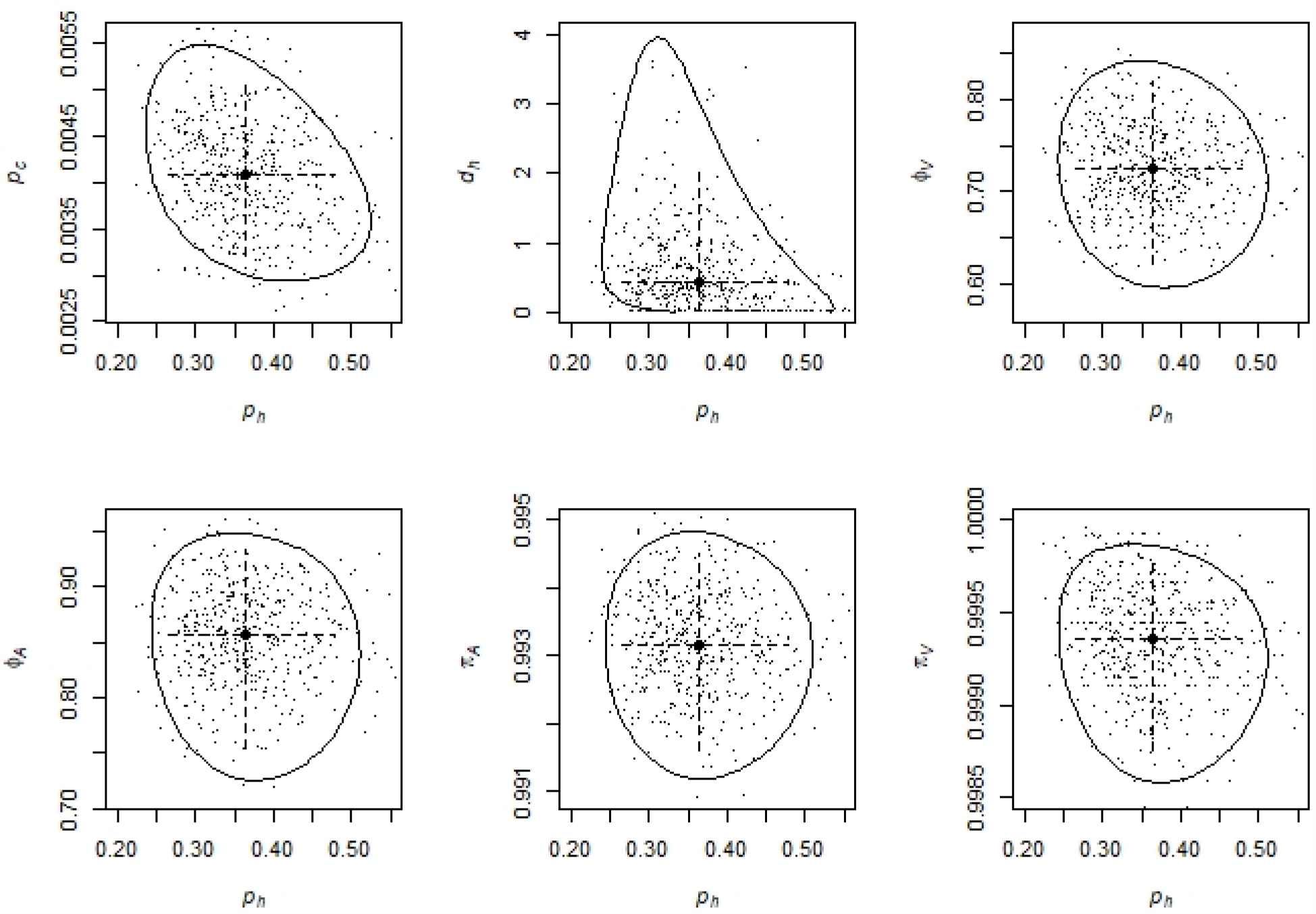
Two-dimension confidence regions for *p*_*h*_ paired with each other parameter Solid curves are the 2-dimensional confidence regions derived from the likelihood ratio test, comparing the likelihood ratio statistic to the 95^th^ percentile of the chi-squared distribution with 2 degrees of freedom. Large solid circle is the MLE estimate and dashed lines are the confidence intervals for each individual parameter derived from the likelihood ratio test (Table 1 main text). Small dots are the MLE estimates from each of 500 simulated data sets generated using parameter values set at the MLE from the actual data (parametric bootstrap).

**Figure S3.**
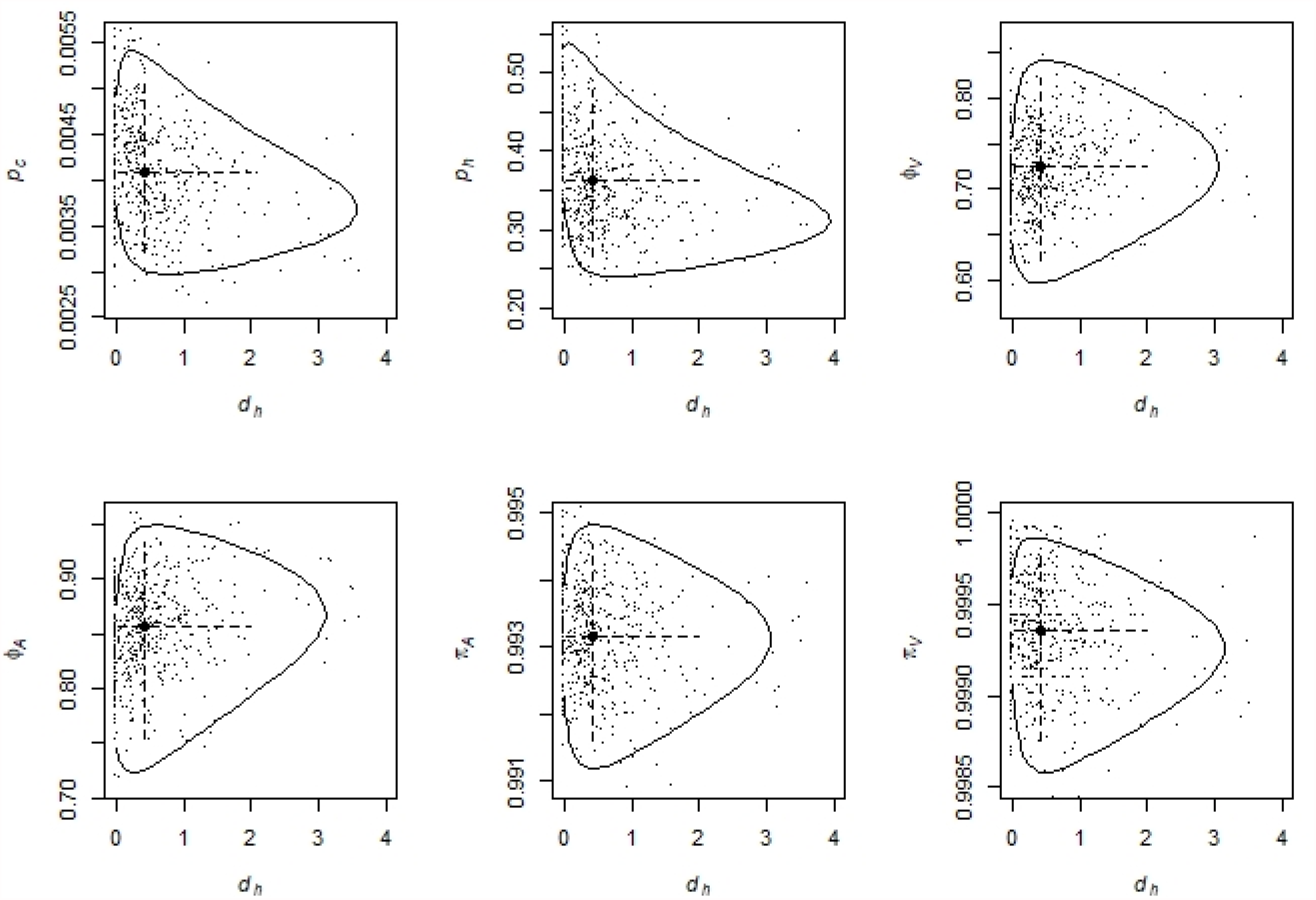
Two-dimension confidence regions for *d*_*h*_ paired with each other parameter Solid curves are the 2-dimensional confidence regions derived from the likelihood ratio test, comparing the likelihood ratio statistic to the 95^th^ percentile of the chi-squared distribution with 2 degrees of freedom. Large solid circle is the MLE estimate and dashed lines are the confidence intervals for each individual parameter derived from the likelihood ratio test (Table 1 main text). Small dots are the MLE estimates from each of 500 simulated data sets generated using parameter values set at the MLE from the actual data (parametric bootstrap).

**Figure S4.**
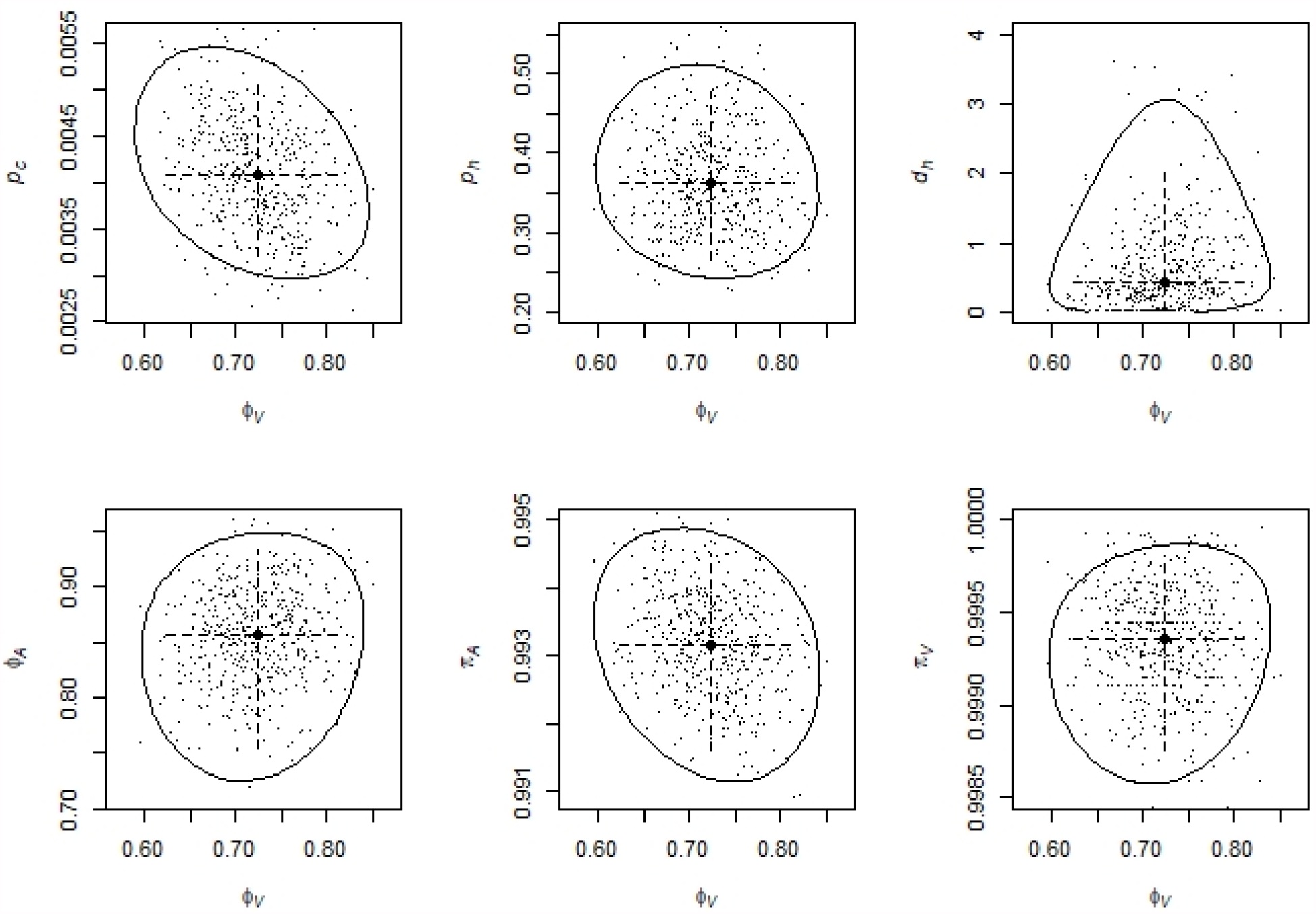
Two-dimension confidence regions for *ϕ*_*V*_ paired with each other parameter Solid curves are the 2-dimensional confidence regions derived from the likelihood ratio test, comparing the likelihood ratio statistic to the 95^th^ percentile of the chi-squared distribution with 2 degrees of freedom. Large solid circle is the MLE estimate and dashed lines are the confidence intervals for each individual parameter derived from the likelihood ratio test (Table 1 main text). Small dots are the MLE estimates from each of 500 simulated data sets generated using parameter values set at the MLE from the actual data (parametric bootstrap).

**Figure S5.**
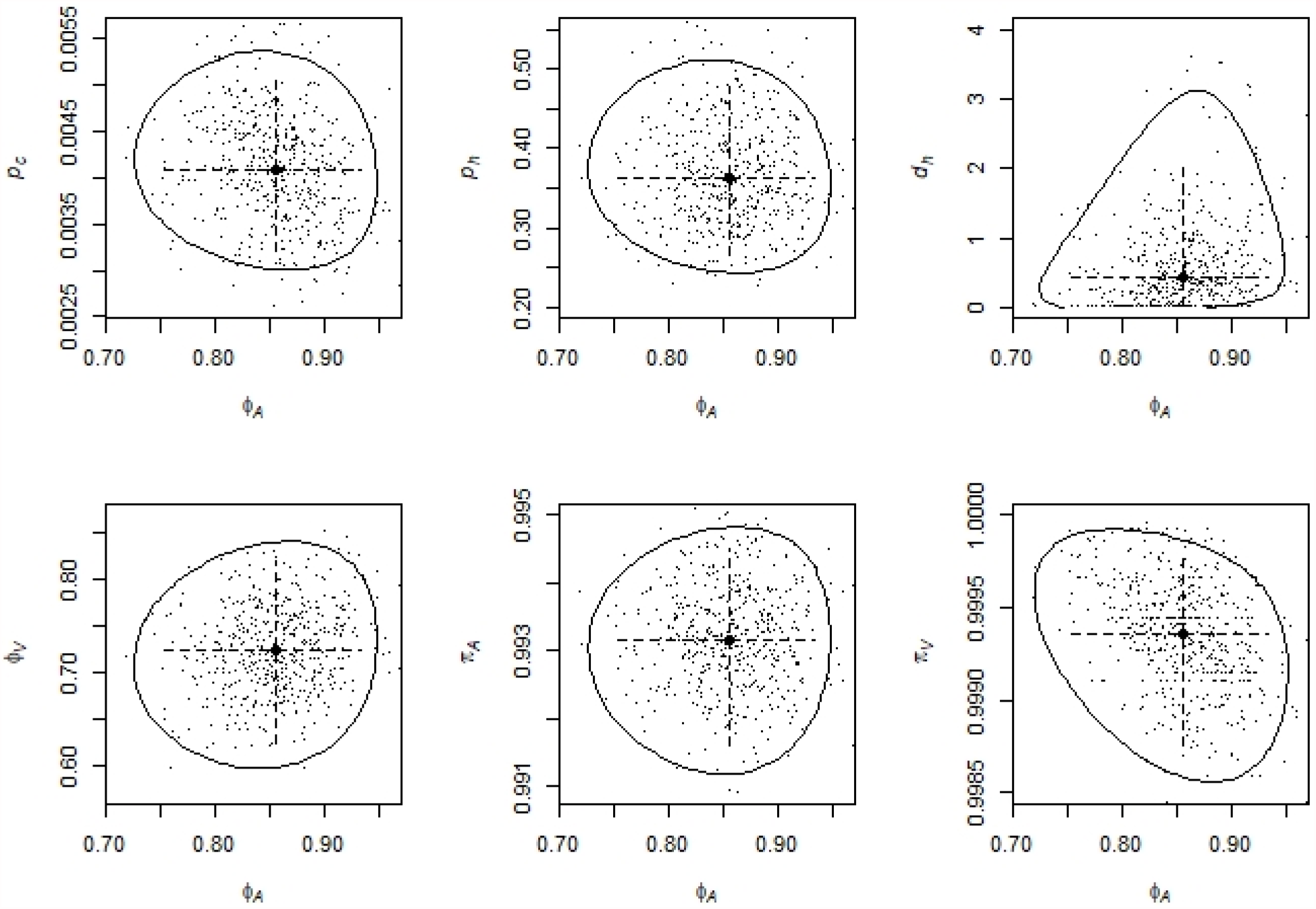
Two-dimension confidence regions for *ϕ*_*A*_ paired with each other parameter Solid curves are the 2-dimensional confidence regions derived from the likelihood ratio test, comparing the likelihood ratio statistic to the 95^th^ percentile of the chi-squared distribution with 2 degrees of freedom. Large solid circle is the MLE estimate and dashed lines are the confidence intervals for each individual parameter derived from the likelihood ratio test (Table 1 main text). Small dots are the MLE estimates from each of 500 simulated data sets generated using parameter values set at the MLE from the actual data (parametric bootstrap).

**Figure S6.**
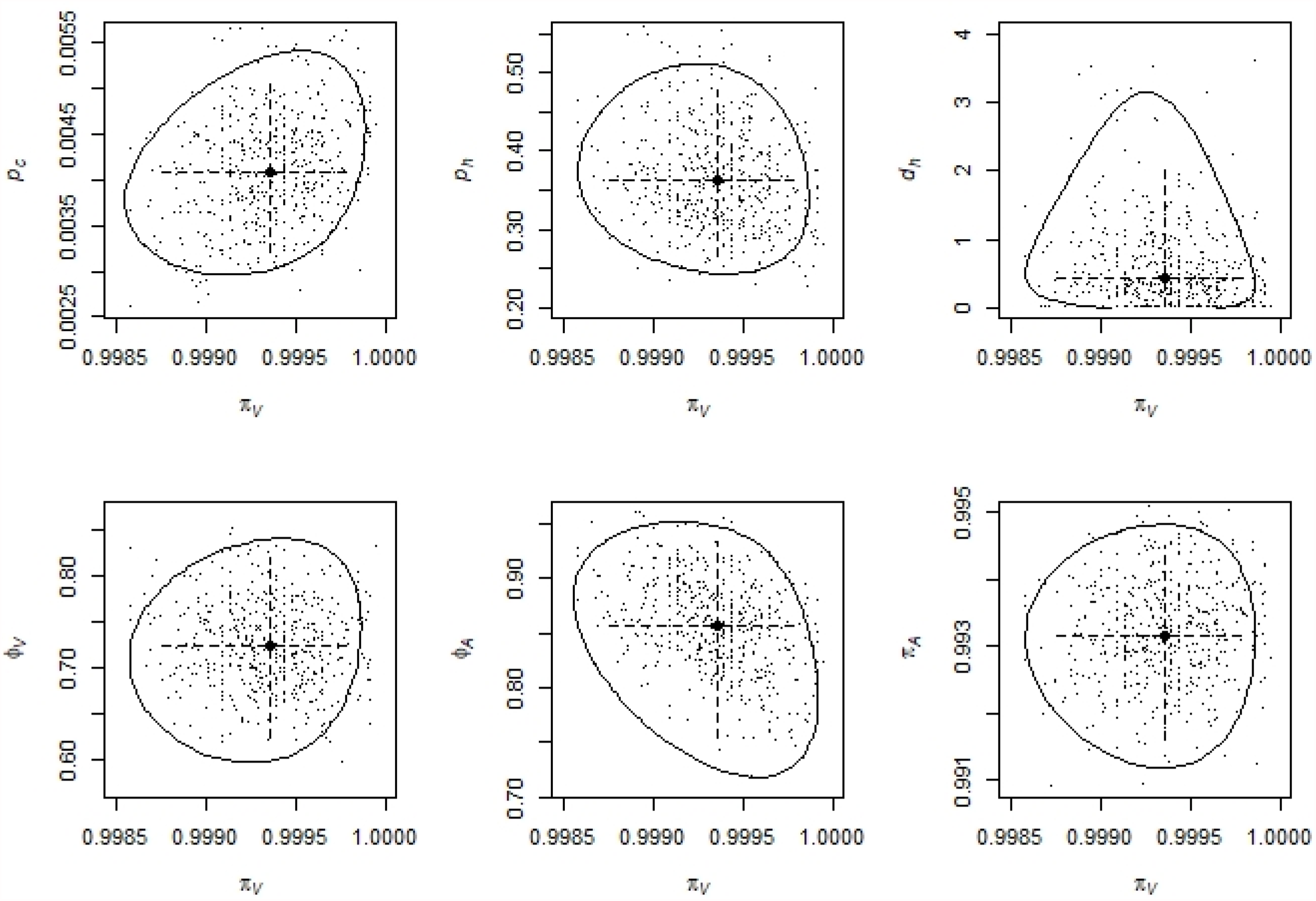
Two-dimension confidence regions for *π*_*V*_ paired with each other parameter Solid curves are the 2-dimensional confidence regions derived from the likelihood ratio test, comparing the likelihood ratio statistic to the 95^th^ percentile of the chi-squared distribution with 2 degrees of freedom. Large solid circle is the MLE estimate and dashed lines are the confidence intervals for each individual parameter derived from the likelihood ratio test (Table 1 main text). Small dots are the MLE estimates from each of 500 simulated data sets generated using parameter values set at the MLE from the actual data (parametric bootstrap).

**Figure S7.**
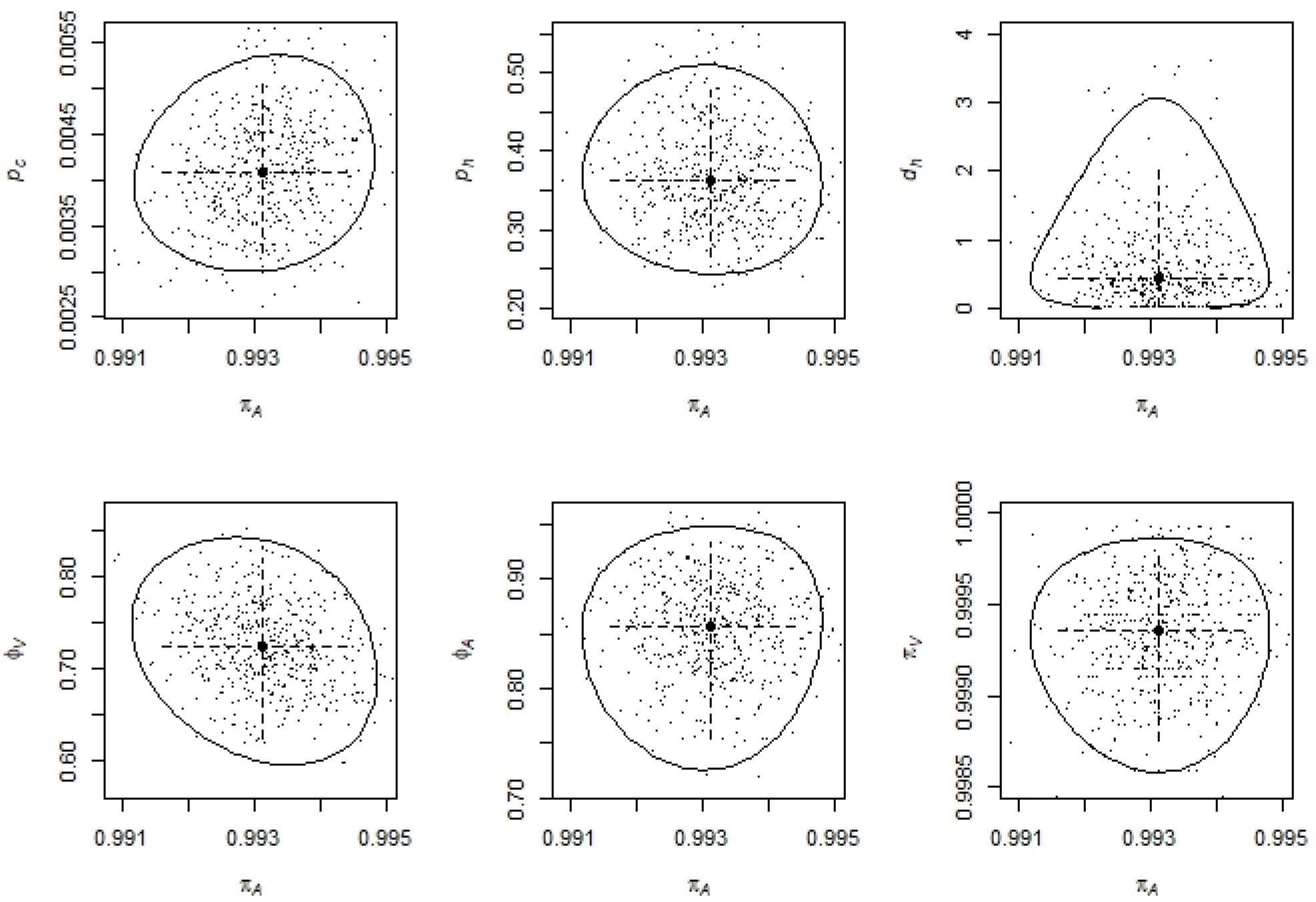
Two-dimension confidence regions for *π*_*A*_ paired with each other parameter Solid curves are the 2-dimensional confidence regions derived from the likelihood ratio test, comparing the likelihood ratio statistic to the 95^th^ percentile of the chi-squared distribution with 2 degrees of freedom. Large solid circle is the MLE estimate and dashed lines are the confidence intervals for each individual parameter derived from the likelihood ratio test (Table 1 main text). Small dots are the MLE estimates from each of 500 simulated data sets generated using parameter values set at the MLE from the actual data (parametric bootstrap).

